# The labour market consequences of extended waiting times for elective inpatient healthcare: an observational study in England using national data

**DOI:** 10.1101/2025.11.21.25340729

**Authors:** Daniel Ayoubkhani, Hannah Aston, Hannah Bunk, Thomas Henstock, Anushka Madhukar, Svetlana Batrakova, Adam Memon, Vahé Nafilyan

## Abstract

**Background:** Waiting times for elective healthcare in England have increased since 2015/16, when the constitutional standard was last met. However, little is known about the economic consequences of extended waits. We estimated the labour market shortfall induced by above-target waiting times for elective hospital admissions in England over a nine-year period.

**Methods:** Cohorts of patients aged 30 to <59 years were identified for 32 treatment specialties using non-emergency inpatient records from NHS hospitals spanning April 2015 to March 2024. Patients were excluded if they had a record within the same treatment specialty from April 2009 to March 2015. Target waiting times were estimated from an exponential distribution with a rate parameter estimated from admissions in 2015/16, assuming patients admitted in each year of the study period remained at their observed percentile of the distribution in that year. Using published trajectories of age-standardised employee pay following elective inpatient treatment, we calculated the additional volume of employment and value of earnings that would be accrued up to five years after the decision to admit, had waiting times remained at target levels.

**Results:** The study population comprised 5.4 million patients (mean age 47.0 years, 54.3% female), of whom 27.3% were treated in multiple specialties during the study period. The biggest decreases in the maximum wait when moving from observed to target waiting times were seen in clinical haematology (13.6 months), medical oncology (12.5 months) and clinical oncology (10.1 months). Losses to earnings due to above-target waiting times were greatest for trauma and orthopaedics services (£375 million over five years), general surgery (£368 million) and urology (£229 million). Losses to employment were largest for trauma and orthopaedics (9,228 person-years of employment over five years), general surgery (5,570 person-years) and pain management (2,631 person-years). Across all 32 analysed treatment specialties, the total five-year loss was estimated at £1.5-2.3 billion for earnings and 20,502-33,130 person-years for employment.

**Conclusions:** Extended waiting times for elective inpatient treatment in England are likely to have impacted the labour market. At the macroeconomic level, reducing waits may lead to increased earnings and economic growth, higher tax receipts and reduced welfare expenditure.

## Introduction

The population of England is living longer than ever before, with life expectancy at birth in 2023 increasing by approximately five years over the past four decades and projected to increase by a further four years over the next half-century^1^. However, these additional years of life are not necessarily being spent in good health^2^. This fall in healthy life expectancy has been mirrored by increased demand for healthcare services in England, with hospital inpatient activity in 2024/25 growing by over 20% compared with 10 years earlier^3,4^, outstripping the 8% population growth over the same period^5^.

The decline in healthy life expectancy and increased demand for healthcare services have coincided with rising levels of economic inactivity; that is, people who are not working nor looking for work. In the final quarter of 2024, 9.3 million working-age people in the UK (over a fifth of the working-age population) were economically inactive, an increase of 8.3% since before the COVID-19 pandemic^6^. Reasons for labour market inactivity include retirement, studying and caring responsibilities, but the group that has grown most rapidly is those who are not working due to long-term ill-health: up by one-third since pre-pandemic, and now representing nearly 30% of total inactivity among working-age people in the UK^6^.

There is a growing body of evidence that chronic health conditions can negatively impact employment capacity, potentially with deleterious consequences for the financial wellbeing of individuals, households and the broader macroeconomy. Research has demonstrated that the onset of physical and mental health conditions could reduce annual earnings by approximately £2,000 for the individuals affected and £1,200 for members of their family^7^.

Incident chronic health conditions, reductions in self-reported general health and onset of disability are also associated with transitions out of employment and into economic inactivity^8^. The economic impacts of disease may be most pronounced in people with a health condition severe enough to require inpatient care. For example, for patients admitted to hospital in England in 2023 alone, the five-year earnings loss has been estimated at a total of £3.7 billion for cardiovascular diseases, £2.9 billion for cancers and £2.4 billion for musculoskeletal conditions^9^.

Elective (non-urgent) procedures for people with chronic health conditions that require inpatient hospital care may be effective at improving labour market engagement and increasing earnings^10^. For example, bariatric surgery has been shown to result in a sustained, long-term increase in the likelihood of paid employment for people with obesity^11^. However, waiting lists and times for elective procedures such as this have grown over the past decade. In 2004, the UK government outlined its aim of all National Health Service (NHS) patients waiting for consultant-led elective care to be treated within 18 weeks of referral^12^, and in 2012, it became a requirement for 92% of patients to be treated within 18 weeks. However, this standard was last met in England in 2015/16, with waiting times increasing steadily since then. In 2024/25, just 61% of patients admitted for elective inpatient treatment had waited less than 18 weeks from referral^13^. Possible reasons for longer waiting times include increased demand for services, and a waitlist backlog caused by service disruption during the COVID-19 pandemic^14^.

There is substantial evidence on the health consequences of prolonged waits for elective inpatient procedures^15–27^. Long waiting times may also have health-mediated economic implications, for example through reduced working hours and productivity, or disengagement from the labour market altogether. However, there is a paucity of empirical evidence on the economic impacts of inpatient waiting times^28,29^. Such evidence could inform government spending decisions in relation to health service delivery, potentially leading to shorter waiting lists and improved patient satisfaction and health outcomes, and ultimately increased labour supply and economic growth; in addition to cutting waiting lists, the latter is a key policy objective of the current UK government^30^.

This study aimed to quantify the total additional earnings and volume of employment accrued over a five-year horizon if waiting times for a wide range of elective inpatient treatments were returned to their 2015/16 levels. The analysis used electronic health records (EHRs) from NHS hospitals in England, with near-whole population coverage, combined with published estimates of average employee pay and the probability of being a paid employee following elective inpatient treatment.

## Methods

### Study design and data

This was a retrospective, observational cohort study of working-age patients admitted to NHS hospitals in England for elective treatment, using NHS Hospital Episode Statistics (HES) Admitted Patient Care (APC) data. One study cohort was identified for each of 32 treatment groups (defined by the specialty in which the consultant was working during the period of care). Collectively, these services covered 96% of elective episodes (excluding childbirth-related activity and diagnostic imaging, which are not treatments for health conditions) among working-age patients recorded in HES APC in 2023/24. Low patient counts in each of the treatment specialties covering the remaining 4% of activity precluded analysis for these specialties.

For each of the 32 treatment specialties, patients were included in the study cohort if they: had a decision-to-admit date during the study period 1 April 2015 to 31 March 2024; had been treated by 31 March 2024; had a non-emergency admission for treatment within the specialty of interest (patients could be included in more than one of the 32 study cohorts); were aged 30 to <59 years at the time of admission (this age range was chosen for consistency with the published treatment effects which we used in conjunction with our analytical outputs, described below); and were enumerated at the 2011 Census of England and Wales (required for record linkage within the analytical environment).

For each specialty, patients were excluded from the study cohort if they had a record for this speciality from 1 April 2009 (the earliest available data) to 31 March 2015. This increased the likelihood that patients included in the study population were being treated for a health condition for the first time within the study period. Patients were also excluded if their admission date or waiting time information was missing or invalid, or if their calculated waiting time exceeded two years (deemed to be the result of erroneous data). In instances where a patient had more than one qualifying record within a treatment specialty, the record with the earliest admission date was kept.

An analytical dataset was curated with one row per person per calendar month. The month containing the admission was assigned ‘Month 0’, and patients were followed for up to 12 months before and 60 months after Month 0. Follow-up was censored when patients were aged <30 years or ≥59 years.

### Calculating observed and target waiting times

Waiting time was calculated as the difference between patients’ decision-to-admit date and admission date. This is not coherent with the definition used in published NHS waiting time statistics^13^ (which is based on time from referral rather than decision-to-admit and accounts for mid-pathway clock-stops). However, referral date is not consistently recorded in the HES APC dataset so could not be used for this analysis.

Each patient was assigned a target waiting time, defined as their expected waiting time had they stayed on the same percentile of the observed waiting time distribution (stratified by treatment specialty and financial year of treatment), but the location and shape of the distribution matched that for patients treated in 2015/16 – the latest year when the target of 92% of patients waiting <18 weeks from referral to treatment was last achieved across consultant-led elective healthcare in England^13^. Candidate statistical distributions for modelling waiting times for admissions in 2015/16 comprised the exponential, gamma, log-normal and Weibull distributions, with the final choice being informed by the Bayesian Information Criterion (BIC).

To align with the estimated treatment effects (see below), which are expressed in terms of calendar months after treatment, the observed and target waiting times were converted from days to calendar months when performing all subsequent calculations.

Observed waiting times being above their target levels during the study period was conceptualised to result in three costs in terms of employment and earnings:

1. Accrual of less treatment effects over the five-year period following decision-to-admit
2. More time spent in the pre-treatment state accruing disbenefit of illness
3. Increased pre-treatment health deterioration means that pay/employment progressed to a lower level following treatment

These components are summarised below and illustrated diagrammatically in **Supplementary Appendix A**.

### Quantifying the loss in treatment effects accrued due to extended waiting times

We used published estimates of the labour market treatment effects, in terms of age-standardised employee earnings (deflated to 2023 prices according to UK consumer price inflation) and person-years of employment, associated with elective inpatient procedures up to 60 months post-treatment^31^. The methodology underpinning these estimates has been detailed elsewhere^10^. We calculated how much treatment effect would have been accrued within an evaluation period of five years from decision-to-admit under target waiting times, compared with what was actually accrued given the observed waiting times over the study period.

### Quantifying the cost of additional exposure to disease due to extended waiting times

We fitted the same segmented regression model as used for the published treatment effects^10^ to estimate the effect of exposure to disease prior to treatment. The model was fitted to the 12 months prior to treatment, with a turning point in the pay/employment trajectory identified during the period to represent disease onset. Thus, the trajectory was divided into several epochs: ‘pre-disease’ (from Month -12 to the breakpoint), ‘pre-treatment’ (from the breakpoint to Month -1), and ‘post-treatment’ (from Month 0 to Month 60). The ‘pre-disease’ trajectory was projected over the ‘pre-treatment’ epoch to provide a counterfactual outcome: an estimate of individuals’ pay/employment had they remained in the ‘pre-disease’ state. When waiting times are above their target levels, patients spent more time in the ‘pre-treatment’ epoch, thus they accrued more disbenefit of illness in terms of their pay/employment.

### Quantifying the cost of pre-treatment health deterioration due to extended waiting times

When waiting times are above target, patients progress further along the pay/employment trajectory in the ‘pre-treatment’ epoch than they otherwise would have done. Therefore, the post-treatment turning point in the trajectory occurs at a lower level of pay/employment (assuming a downward pre-treatment trajectory), and this difference in level was assumed to be constant and to persist throughout the post-treatment period.

### Total labour market cost of extended waiting times

For each treatment specialty, the cumulative (across all patients and months) loss to pay/employment due to above-target waiting times over the five-year evaluation period following decision-to-admit was calculated as the sum of: the difference in the cumulative treatment effect under target and observed waiting times; the effect of accruing more disbenefit of disease; the effect of more pre-treatment health deterioration on post-treatment levels.

As the 32 study cohorts are not mutually exclusive, some of the estimated loss in pay/employment due to above-target waiting times for a given treatment specialty may be attributable to patients being treated in other specialities. Thus, there will be some degree of double-counting if estimates of the labour market costs of extended waiting times are simply summed across treatment specialties, giving an upper bound on the estimated total loss due to above-target waits. To provide a more conservative estimate, cohort sizes were re-calculated under the assumption that patients belonging to multiple study cohorts experienced losses in pay/employment due to above-target waiting times for just one specialty in which they were treated: the one with the smallest per-person effect size. The re-calculated cohort sizes were then multiplied by the per-person effect sizes and summed across the 32 treatment specialties, giving a lower bound on the estimated total loss due to above-target waits.

For each treatment specialty, 95% confidence intervals (CIs) were constructed around the point estimates of the change in the cumulative treatment effect under observed and target waiting times by assuming that this quantity follows a normal distribution.

Data preparation was conducted using Sparklyr version 1.9 and statistical analyses were performed using R version 4.4. Further details of the statistical methodology can be found in **Supplementary Appendix A**.

## Results

### Description of the study population

The study population comprised 5,363,712 patients (mean age 47.0 years, 54.3% female out of *n*=5,359,033 with a known sex), derived as per **Figure 1**. During the study period, 72.7% of patients were treated in one specialty, 20.0% in two specialties and 5.4% in ≥3 specialties. The 32 treatment-specific cohorts ranged from *n*=11,516 for the anaesthetic service to *n*=1,615,833 for gastroenterology (**Table 1**). The study cohorts were generally older and more likely to be female than the general population of England aged 30 to <59 years (mean age 44.0 years, 51.1% female in 2021^32^).

**Table 1.**
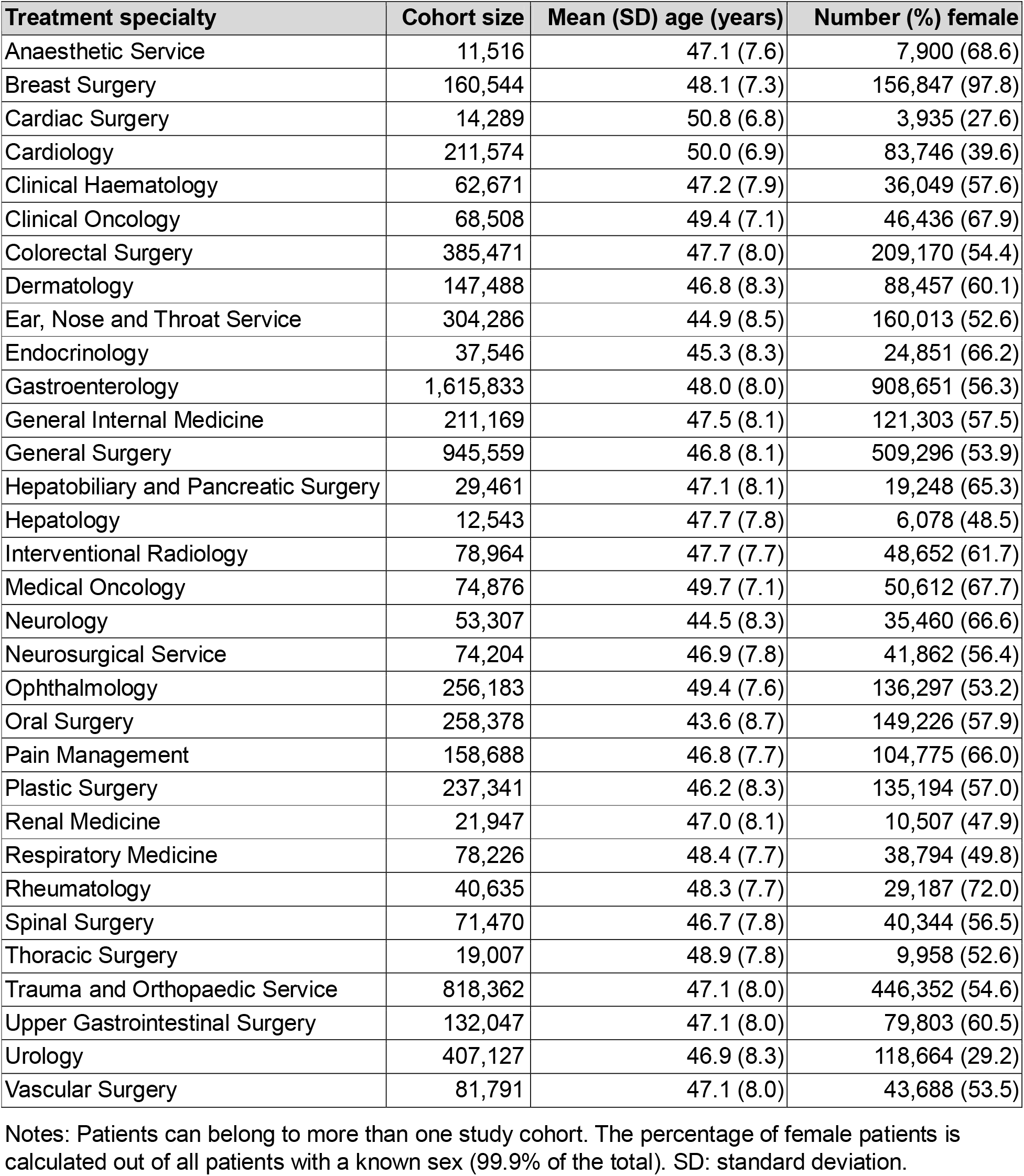
Age and sex distributions of the study cohorts at time of treatment.

| Treatment specialty | Cohort size | Mean (SD) age (years) | Number (%) female |
| --- | --- | --- | --- |
| Anaesthetic Service | 11,516 | 47.1 (7.6) | 7,900 (68.6) |
| Breast Surgery | 160,544 | 48.1 (7.3) | 156,847 (97.8) |
| Cardiac Surgery | 14,289 | 50.8 (6.8) | 3,935 (27.6) |
| Cardiology | 211,574 | 50.0 (6.9) | 83,746 (39.6) |
| Clinical Haematology | 62,671 | 47.2 (7.9) | 36,049 (57.6) |
| Clinical Oncology | 68,508 | 49.4 (7.1) | 46,436 (67.9) |
| Colorectal Surgery | 385,471 | 47.7 (8.0) | 209,170 (54.4) |
| Dermatology | 147,488 | 46.8 (8.3) | 88,457 (60.1) |
| Ear, Nose and Throat Service | 304,286 | 44.9 (8.5) | 160,013 (52.6) |
| Endocrinology | 37,546 | 45.3 (8.3) | 24,851 (66.2) |
| Gastroenterology | 1,615,833 | 48.0 (8.0) | 908,651 (56.3) |
| General Internal Medicine | 211,169 | 47.5 (8.1) | 121,303 (57.5) |
| General Surgery | 945,559 | 46.8 (8.1) | 509,296 (53.9) |
| Hepatobiliary and Pancreatic Surgery | 29,461 | 47.1 (8.1) | 19,248 (65.3) |
| Hepatology | 12,543 | 47.7 (7.8) | 6,078 (48.5) |
| Interventional Radiology | 78,964 | 47.7 (7.7) | 48,652 (61.7) |
| Medical Oncology | 74,876 | 49.7 (7.1) | 50,612 (67.7) |
| Neurology | 53,307 | 44.5 (8.3) | 35,460 (66.6) |
| Neurosurgical Service | 74,204 | 46.9 (7.8) | 41,862 (56.4) |
| Ophthalmology | 256,183 | 49.4 (7.6) | 136,297 (53.2) |
| Oral Surgery | 258,378 | 43.6 (8.7) | 149,226 (57.9) |
| Pain Management | 158,688 | 46.8 (7.7) | 104,775 (66.0) |
| Plastic Surgery | 237,341 | 46.2 (8.3) | 135,194 (57.0) |
| Renal Medicine | 21,947 | 47.0 (8.1) | 10,507 (47.9) |
| Respiratory Medicine | 78,226 | 48.4 (7.7) | 38,794 (49.8) |
| Rheumatology | 40,635 | 48.3 (7.7) | 29,187 (72.0) |
| Spinal Surgery | 71,470 | 46.7 (7.8) | 40,344 (56.5) |
| Thoracic Surgery | 19,007 | 48.9 (7.8) | 9,958 (52.6) |
| Trauma and Orthopaedic Service | 818,362 | 47.1 (8.0) | 446,352 (54.6) |
| Upper Gastrointestinal Surgery | 132,047 | 47.1 (8.0) | 79,803 (60.5) |
| Urology | 407,127 | 46.9 (8.3) | 118,664 (29.2) |
| Vascular Surgery | 81,791 | 47.1 (8.0) | 43,688 (53.5) |

Notes: Patients can belong to more than one study cohort. The percentage of female patients is calculated out of all patients with a known sex (99.9% of the total). SD: standard deviation.

**Figure 1.**
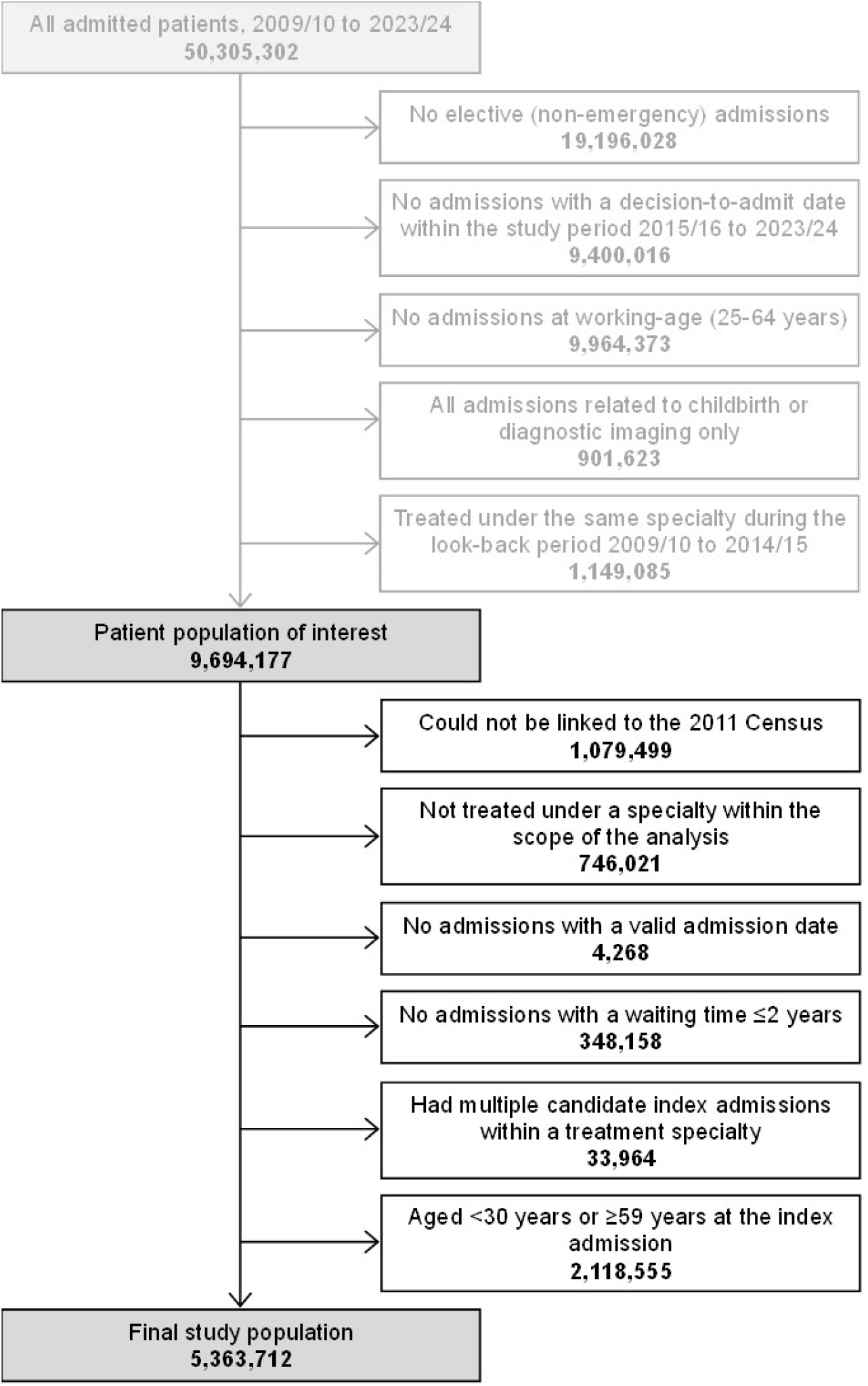
Study population flow diagram Notes: For this analysis, the working-age population was defined as individuals aged 25-64 years, thus omitting most people in full-time education. Admissions likely to be related to childbirth were defined as those under the following treatment specialties: obstetrics, fetal medicine, midwifery, and the community sexual and reproductive health service. Each patient’s ‘index admission’ was their first study-eligible admission within the study period under each treatment specialty.

### Differences between observed and target waiting times

For modelling waiting times for admissions in 2015/16, no single statistical distribution consistently minimised the BIC across the 32 treatment specialties (**Supplementary Table 1**). Therefore, the most parsimonious specification (the exponential distribution) was chosen for the main analysis. The gamma distribution was used in a sensitivity analysis as this specification is a generalisation of the exponential distribution; or conversely, the exponential distribution is a special case of the gamma distribution.

The exponential probability density function provided a reasonable representation of the waiting time distribution in 2015/16 for most treatment specialties (**Supplementary Figure 1**), but with a tendency to over-estimate the proportion of patients with waiting times ≤7 days (this group may include emergency admissions that were not coded as such in the HES dataset), which was not remedied by the more flexible gamma distribution (**Supplementary Figure 2**).

Across the full study period April 2015 to March 2024, the biggest differences between median observed and target waiting times were for pain management (from 60 to 42 days), cardiac surgery (from 59 to 41 days) and vascular surgery (from 59 to 41 days) (**Figure 2**). The biggest decreases in the maximum wait when moving from observed to target waiting times were seen in clinical haematology (from 23.4 to 9.8 months), medical oncology (from 23.4 to 10.9 months) and clinical oncology (from 24.0 to 13.9 months).

**Figure 2.**
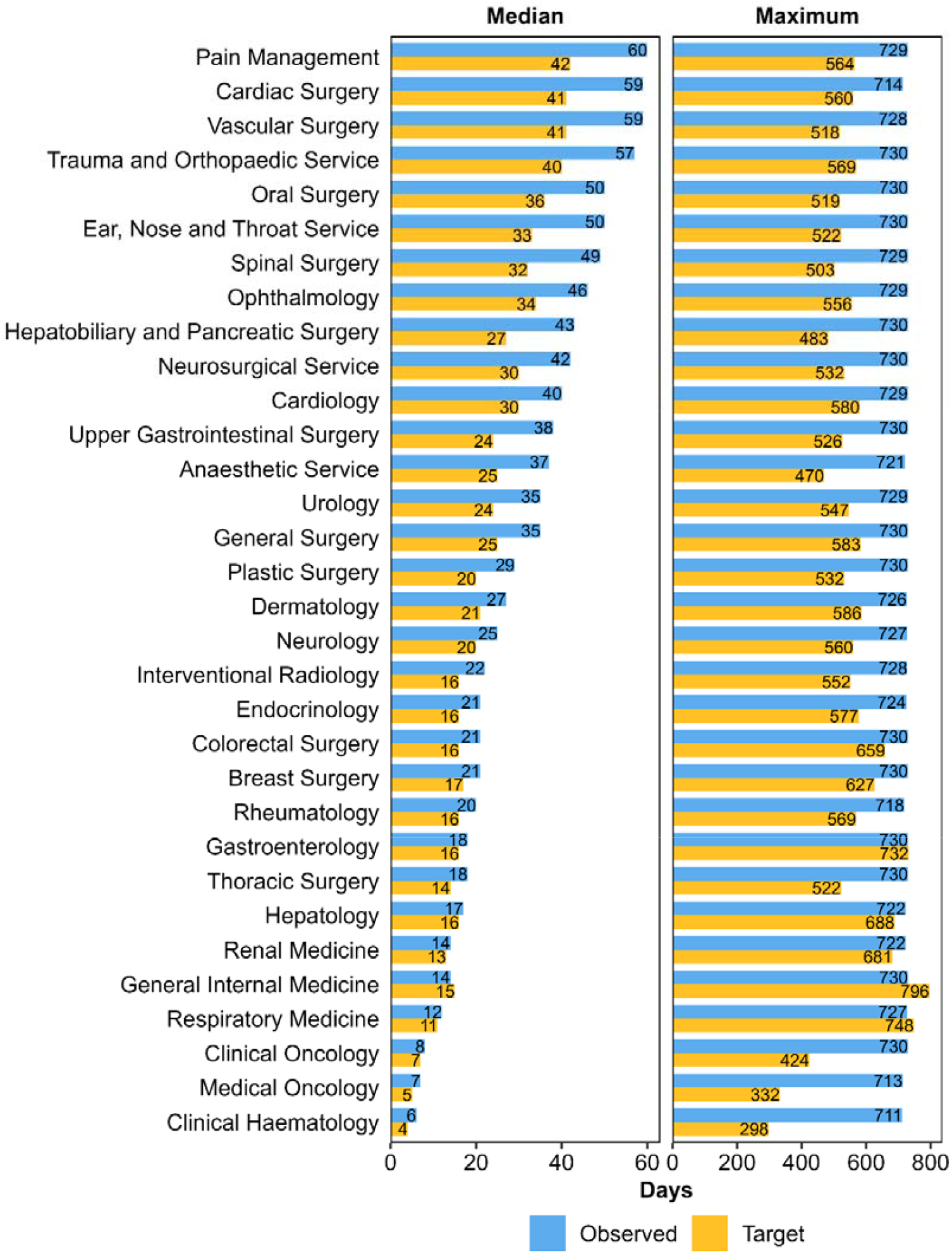
Median and maximum observed and target waiting times Notes: Median and maximum waiting times were calculated among patients with a decision-to-admit date between 1 April 2015 and 31 March 2024 who were treated by 31 March 2024. Patients were excluded from the study population if their calculated time from decision-to-admit to admission exceeded two years (deemed to be the result of erroneous data), hence the maximum observed wait is 730 days. Each patient’s target waiting time was defined as their expected waiting time had they stayed on the same percentile of the observed waiting time distribution (modelled as an exponential distribution, stratified by treatment specialty and financial year of treatment), but the rate parameter of the distribution (which defines it location and shape) matched that for patients treated in 2015/16.

### Labour market impacts of above-target waiting times

Among patients who received a decision to admit and were treated during the nine-year study period, losses to earnings accrued over the five years post-treatment due to above-target waiting times were greatest for the trauma and orthopaedics service (£375 million, 95% CI: £373 million to £376 million), general surgery (£368 million, £367 million to £370 million) and urology (£229 million, £228 million to £230 million) (**Table 2**). Five-year losses to employment due to above-target waiting times were greatest for the trauma and orthopaedics service (9,228 person-years of employment, 95% CI: 9,201 to 9,256 person-years), general surgery (5,570 person-years, 5,551 to 5,590 person-years) and pain management (2,631 person-years, 2,612 to 2,651 person-years).

**Table 2.**
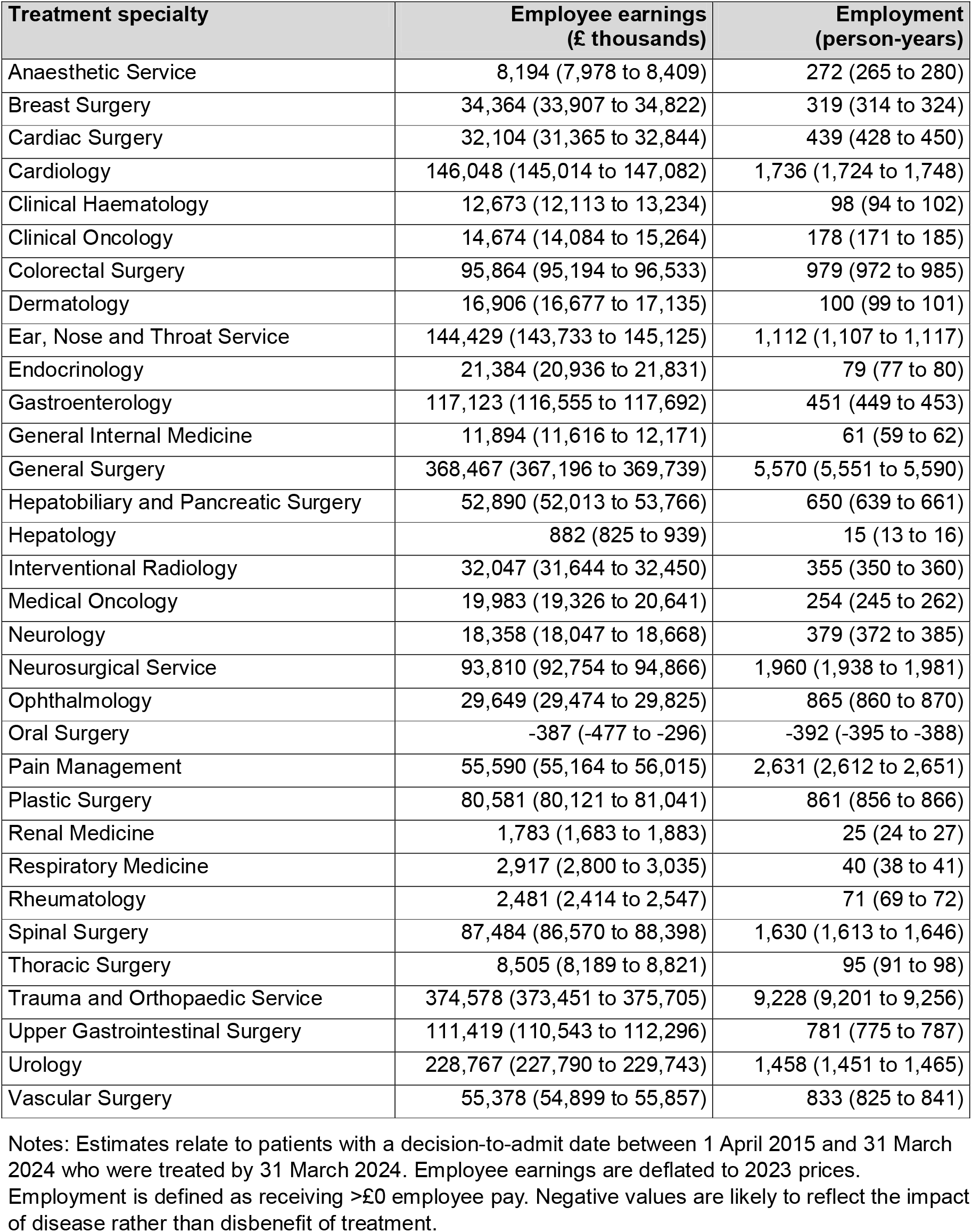
Estimated labour market impacts (point estimates and 95% confidence intervals) of above-target waiting times over the study period, accrued over five years post-admission.

| Treatment specialty | Employee earnings (£ thousands) | Employment (person-years) |
| --- | --- | --- |
| Anaesthetic Service | 8,194 (7,978 to 8,409) | 272 (265 to 280) |
| Breast Surgery | 34,364 (33,907 to 34,822) | 319 (314 to 324) |
| Cardiac Surgery | 32,104 (31,365 to 32,844) | 439 (428 to 450) |
| Cardiology | 146,048 (145,014 to 147,082) | 1,736 (1,724 to 1,748) |
| Clinical Haematology | 12,673 (12,113 to 13,234) | 98 (94 to 102) |
| Clinical Oncology | 14,674 (14,084 to 15,264) | 178 (171 to 185) |
| Colorectal Surgery | 95,864 (95,194 to 96,533) | 979 (972 to 985) |
| Dermatology | 16,906 (16,677 to 17,135) | 100 (99 to 101) |
| Ear, Nose and Throat Service | 144,429 (143,733 to 145,125) | 1,112 (1,107 to 1,117) |
| Endocrinology | 21,384 (20,936 to 21,831) | 79 (77 to 80) |
| Gastroenterology | 117,123 (116,555 to 117,692) | 451 (449 to 453) |
| General Internal Medicine | 11,894 (11,616 to 12,171) | 61 (59 to 62) |
| General Surgery | 368,467 (367,196 to 369,739) | 5,570 (5,551 to 5,590) |
| Hepatobiliary and Pancreatic Surgery | 52,890 (52,013 to 53,766) | 650 (639 to 661) |
| Hepatology | 882 (825 to 939) | 15 (13 to 16) |
| Interventional Radiology | 32,047 (31,644 to 32,450) | 355 (350 to 360) |
| Medical Oncology | 19,983 (19,326 to 20,641) | 254 (245 to 262) |
| Neurology | 18,358 (18,047 to 18,668) | 379 (372 to 385) |
| Neurosurgical Service | 93,810 (92,754 to 94,866) | 1,960 (1,938 to 1,981) |
| Ophthalmology | 29,649 (29,474 to 29,825) | 865 (860 to 870) |
| Oral Surgery | -387 (-477 to -296) | -392 (-395 to -388) |
| Pain Management | 55,590 (55,164 to 56,015) | 2,631 (2,612 to 2,651) |
| Plastic Surgery | 80,581 (80,121 to 81,041) | 861 (856 to 866) |
| Renal Medicine | 1,783 (1,683 to 1,883) | 25 (24 to 27) |
| Respiratory Medicine | 2,917 (2,800 to 3,035) | 40 (38 to 41) |
| Rheumatology | 2,481 (2,414 to 2,547) | 71 (69 to 72) |
| Spinal Surgery | 87,484 (86,570 to 88,398) | 1,630 (1,613 to 1,646) |
| Thoracic Surgery | 8,505 (8,189 to 8,821) | 95 (91 to 98) |
| Trauma and Orthopaedic Service | 374,578 (373,451 to 375,705) | 9,228 (9,201 to 9,256) |
| Upper Gastrointestinal Surgery | 111,419 (110,543 to 112,296) | 781 (775 to 787) |
| Urology | 228,767 (227,790 to 229,743) | 1,458 (1,451 to 1,465) |
| Vascular Surgery | 55,378 (54,899 to 55,857) | 833 (825 to 841) |

Notes: Estimates relate to patients with a decision-to-admit date between 1 April 2015 and 31 March 2024 who were treated by 31 March 2024. Employee earnings are deflated to 2023 prices. Employment is defined as receiving >£0 employee pay. Negative values are likely to reflect the impact of disease rather than disbenefit of treatment.

Across all 32 analysed treatment specialties and ignoring any double-counting of patients between treatment specialties, the five-year loss due to above-target waiting times summed to £2,281 million (95% CI: £2,264 million to £2,289 million) for earnings and 33,130 person-years (32,887 to 33,374 person-years) for employment. These totals represent upper bounds on the estimated total impact of extended waiting times.

After adjusting the cohort sizes to account for some patients being treated under multiple specialties and multiplying by the estimated per-person effect sizes, the total five-year loss across all 32 specialties was conservatively estimated at £1,470 million (95% CI: £1,460 million to £1,480 million) for earnings and 20,502 person-years (20,363 to 20,641 person-years) for employment. These totals represent lower bounds on the estimated total impact of extended waiting times.

In the sensitivity analysis, very similar aggregate results were obtained when using the gamma distribution rather than the exponential distribution to infer target waiting times, with the total five-year loss conservatively estimated to be £1,489 million (95% CI: £1,478 million to £1,501 million) for earnings and 20,534 person-years (20,381 to 20,687 person-years) for employment. Estimates for individual treatment specialties based on the gamma distribution can be found in **Supplementary Table 2**.

## Discussion

Among all elective inpatients with a decision-to-admit between April 2015 and March 2024 who were treated in NHS hospitals in England, the total five-year loss to employee earnings due to above-target waiting times ranged from £1.5 billion to £2.3 billion. The total five-year loss to payrolled employment ranged from 20,502 to 33,130 person-years. The effect on earnings is likely to be at least partly driven by the effect on employment, in addition to other factors such as reduced working hours and workplace performance.

This study was not intended to be a cost-benefit analysis of policies aimed at reducing waiting times for elective care. However, to contextualise our finding that approximately £2 billion in additional earnings could have been generated if waiting times had remained at the constitutional target, this figure represents less than 0.2% of the total amount spent on healthcare in England over the study period (excluding expenditure directly related to the COVID-19 pandemic)^33^.

There is substantial evidence on the health consequences of prolonged waits for elective health interventions, with much of the research for inpatients focussing on orthopaedic surgery^15–25^ as well as prostatectomy^15^, coronary bypass^26^ and treatments for varicose veins^24,27^, hernia^24,27^ and gallstones^27^. The results of these studies are mixed, with some but not all finding longer waits to be associated with poorer health and quality-of-life outcomes (possibly reflecting the small samples sizes and specific patient populations studied). Above-target waits for elective treatments have also been found to be associated with increased healthcare utilisation^34^, demonstrating one potential source of the economic cost of delayed treatment. However, to our knowledge, this is the first study to use individual-level, longitudinal data to explore the labour market consequences of extended waiting times for elective inpatient procedures in England.

One study using administrative data from Norway found increased waits for orthopaedic surgery to be associated with reduced earnings and increased sickness absence^28^, but it is unclear whether these results can be generalised to other countries and treatments. A study in England attempted to quantify the economic benefit of reducing elective waiting times^29^, but this was largely based on aggregate rather than record-level data and was in relation to a specific NHS delivery plan^35^ which is now outdated. Studies in the UK^36^ and the Netherlands^37^ have found links between waiting times for mental health treatment and subsequent labour market outcomes, but in relation to specialised, community-based services rather than hospital inpatient settings, which is the focus of the present study.

### Strengths and limitations

The main strength of this study is its use of EHRs covering all NHS hospitals in England, thus providing a large sample size (5.4 million patients in total) with whole-population coverage. This meant that results could be produced for individual treatment specialties (collectively covering the vast majority of relevant elective activity) and that study estimates can safely be generalized to the population of interest.

The limitations of this study mainly stem from those of the treatment effects on which it draws, all of which imply that our results are likely to be underestimates of the total economic cost of extended waiting times. The study population only included people aged 30-59 years rather than the broader working-age population (typically defined as those aged 16-64 years in the UK^38^). The treatment effects only capture the contribution of payrolled employees, not the self-employed (who make up 13% of the UK workforce^6^). The estimated treatment effects, and thus our estimates of the impact of extended waiting times, do not vary by time waited for treatment. There is evidence of an inverse relationship between treatment effects and waiting time^28^, perhaps because timely treatment may achieve a bigger effect for progressive health conditions and potentially means less time being detached from the labour market. The treatment effects and our study estimates also do not capture impacts to the economy beyond those experienced by individuals, such as impacts on output/productivity within firms, welfare benefit payments and healthcare utilisation costs.

The HES dataset only includes records for patients treated during the study period; those who were still waiting for treatment at the end of the period are not present on the dataset. Therefore, there will be a degree of right-censoring inherent in our analysis and the cohort sizes will be smaller than the true number of patients with a decision-to-admit during the study period, so the aggregate effect of above-target waiting times will be underestimated.

When estimating patients’ target waiting times, the exponential distribution generally provided a reasonable fit to the waiting time distribution in 2015/16, except for patients who waited a week or less for treatment. This local lack of fit at the lower end of the waiting time distribution is unlikely to have affected the findings of the analysis, as patients with the longest waiting times accrue the most disbenefit of extended waits and therefore contribute most to our estimates.

Confidence intervals around estimates of the impact of extended waiting times do not reflect uncertainty inherent in the input values to the calculations (age-standardised trajectories, modelled treatment and exposure-to-disease effects, and expected waiting times under the target distribution). These are treated as known quantities rather than statistical estimates, so the final confidence intervals are likely to underestimate total uncertainty.

## Conclusions

Lengthening waiting times for elective inpatient treatment in England since 2015/16 are likely to have had non-negligible consequences for the labour market, including multi-billion-pound losses to aggregate earnings. This research adds to the growing evidence base on the relationship between population health and the economy, demonstrating that reducing health service waiting times may lead to increased earnings for individuals and their households. At the macroeconomic level, shorter waits may contribute to higher tax receipts, reduced welfare expenditure and, crucially, more economic growth.

The results of this study can be used in economic appraisals of health service and policy interventions aimed at reducing waiting lists for planned inpatient procedures. Indeed, based on this research and estimates of future healthcare demand, the NHS has projected that if waiting times for elective hospital procedures returned to the constitutional standard (92% of patients treated within 18 weeks of referral) by 2030/31, employee earnings in England would increase cumulatively by £2.7 billion over the period and an addition 63,000 person-years of employment would be accrued^39^.

Further work is needed to investigate the extent to which the labour market outcomes of health interventions are modified by time waited for treatment, which could then be incorporated into refined estimates of the impact of reduced waiting times on aggregate earnings and employment. Also required is research into the labour market consequences of extended waiting times for elective treatments administered outside of inpatient settings, such as outpatient and community services, which were beyond the scope of this study.

## Abbreviations

BIC: Bayesian Information Criterion
CI: Confidence interval
HER: Electronic health record
HES APC: Hospital Episode Statistics – Admitted Patient Care
NHS: National Health Service
SD: Standard deviation

## Declarations

### Ethics approval and consent to participate

Ethical approval for this work was obtained from the National Statistician’s Data Ethics Advisory Committee (NSDEC23(18)) according to the ethical principles outlined here: https://uksa.statisticsauthority.gov.uk/the-authority-board/committees/national-statisticians-advisory-committees-and-panels/national-statisticians-data-ethics-advisory-committee/ethical-principles/

The study also conforms to the World Medical Association’s Declaration of Helsinki – Ethical Principles for Medical Research Involving Human Participants: https://www.wma.net/policies-post/wma-declaration-of-helsinki/

Individual consent to participate in the study was not required for this analysis of administrative data. Sections 45B and 45C of the Statistics and Registration Service Act (SRSA) 2007 provide a power for Crown bodies and other public authorities (respectively) to share the data they hold with the Office for National Statistics (ONS) to produce statistics, without requiring the consent of the individuals to whom the data relate. Although General Data Protection Regulation (GDPR) usually restricts the use of personal data to the purpose for which the data were originally collected, there is special provision within the UK GDPR that permits further use of data for statistical and research purposes (Article 6(1)(e) of the UK GDPR). Further information can be found in the ONS’s health data policy: https://www.ons.gov.uk/aboutus/transparencyandgovernance/datastrategy/datapolicies/collectingandusinghealthdata

### Consent for publication

Not applicable.

### Data availability

Researchers can apply to access Hospital Episode Statistics data via NHS England’s Data Access Request Service (DARS): https://digital.nhs.uk/services/data-access-request-service-dars

### Authors’ contributions

DA and VN designed the study. DA, HA and HB performed the data analysis. All authors contributed to interpretation of the study results. DA wrote the first draft of the manuscript. All authors reviewed and revised the manuscript and approved the final version.

## Acknowledgements

The authors would like to thank David Goll at Number 10 Data Science, and Chris Worsfold and Henry Foster at the Department of Health and Social Care, for their feedback on the study design. We are also grateful to the following national clinical directors and specialty advisors at NHS England for their valuable insights when interpreting the study results: Dawn Adamson, Mark Cheetham, Jeremy Davis, Jonathan Fuld, Harriet Gordon, Martin Heaton, Tracey Irvine, Lesley Kay, Sue Mann, John McGrath, Adam Millican-Slater, Niranjanan Nirmalananthan, Nick Phillips, Simon Ray, Colin Rees, Alexandra Severns, Doug West.

---

**Supplementary Table 1.**
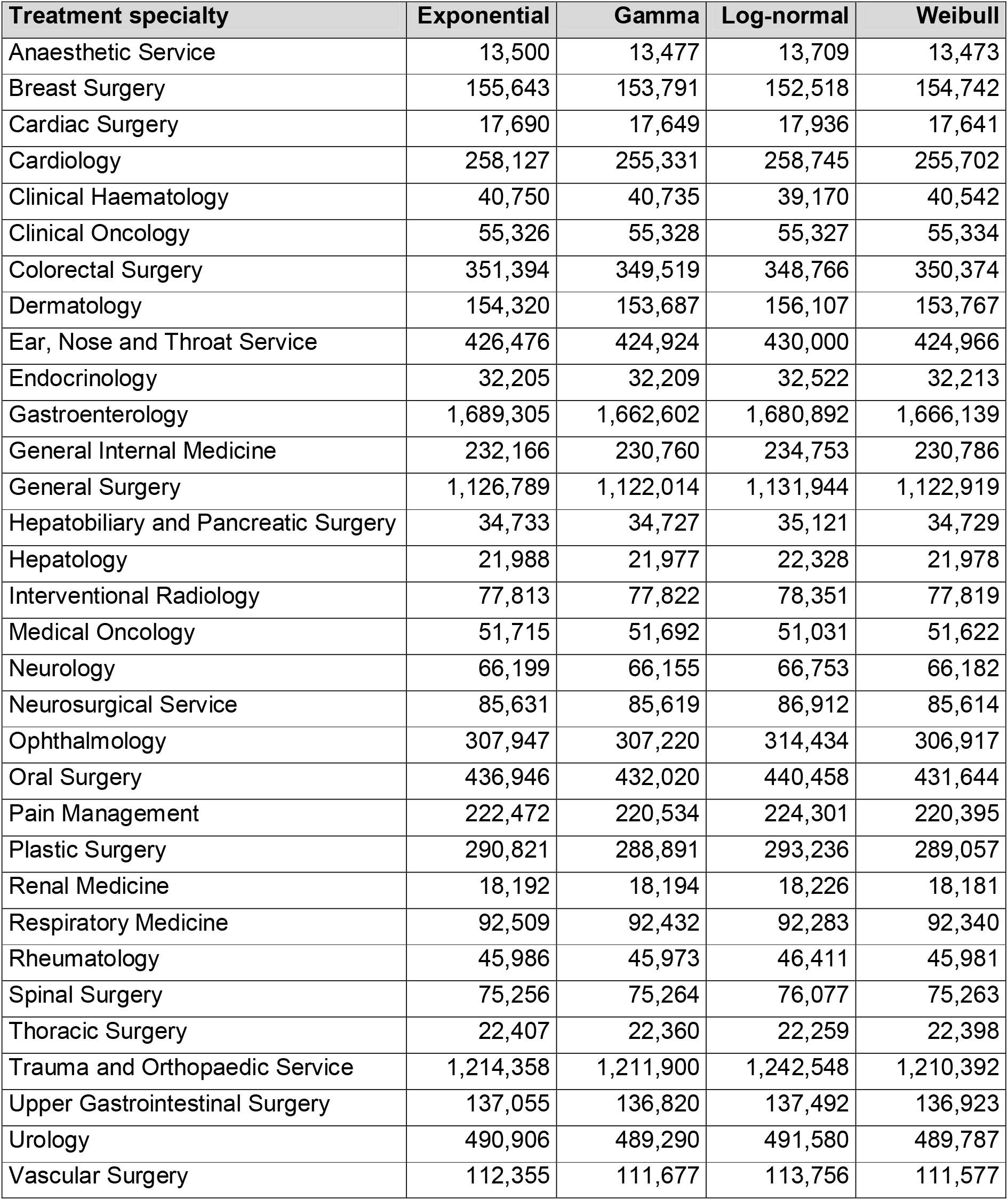
Bayesian Information Criterion (BIC) values for modelling the distribution of waiting times for admissions in 2015/16.

| **Treatment specialty** | **Exponential** | **Gamma** | **Log-normal** | **Weibull** |
| --- | --- | --- | --- | --- |
| Anaesthetic Service | 13,500 | 13,477 | 13,709 | 13,473 |
| Breast Surgery | 155,643 | 153,791 | 152,518 | 154,742 |
| Cardiac Surgery | 17,690 | 17,649 | 17,936 | 17,641 |
| Cardiology | 258,127 | 255,331 | 258,745 | 255,702 |
| Clinical Haematology | 40,750 | 40,735 | 39,170 | 40,542 |
| Clinical Oncology | 55,326 | 55,328 | 55,327 | 55,334 |
| Colorectal Surgery | 351,394 | 349,519 | 348,766 | 350,374 |
| Dermatology | 154,320 | 153,687 | 156,107 | 153,767 |
| Ear, Nose and Throat Service | 426,476 | 424,924 | 430,000 | 424,966 |
| Endocrinology | 32,205 | 32,209 | 32,522 | 32,213 |
| Gastroenterology | 1,689,305 | 1,662,602 | 1,680,892 | 1,666,139 |
| General Internal Medicine | 232,166 | 230,760 | 234,753 | 230,786 |
| General Surgery | 1,126,789 | 1,122,014 | 1,131,944 | 1,122,919 |
| Hepatobiliary and Pancreatic Surgery | 34,733 | 34,727 | 35,121 | 34,729 |
| Hepatology | 21,988 | 21,977 | 22,328 | 21,978 |
| Interventional Radiology | 77,813 | 77,822 | 78,351 | 77,819 |
| Medical Oncology | 51,715 | 51,692 | 51,031 | 51,622 |
| Neurology | 66,199 | 66,155 | 66,753 | 66,182 |
| Neurosurgical Service | 85,631 | 85,619 | 86,912 | 85,614 |
| Ophthalmology | 307,947 | 307,220 | 314,434 | 306,917 |
| Oral Surgery | 436,946 | 432,020 | 440,458 | 431,644 |
| Pain Management | 222,472 | 220,534 | 224,301 | 220,395 |
| Plastic Surgery | 290,821 | 288,891 | 293,236 | 289,057 |
| Renal Medicine | 18,192 | 18,194 | 18,226 | 18,181 |
| Respiratory Medicine | 92,509 | 92,432 | 92,283 | 92,340 |
| Rheumatology | 45,986 | 45,973 | 46,411 | 45,981 |
| Spinal Surgery | 75,256 | 75,264 | 76,077 | 75,263 |
| Thoracic Surgery | 22,407 | 22,360 | 22,259 | 22,398 |
| Trauma and Orthopaedic Service | 1,214,358 | 1,211,900 | 1,242,548 | 1,210,392 |
| Upper Gastrointestinal Surgery | 137,055 | 136,820 | 137,492 | 136,923 |
| Urology | 490,906 | 489,290 | 491,580 | 489,787 |
| Vascular Surgery | 112,355 | 111,677 | 113,756 | 111,577 |

**Supplementary Table 2.**
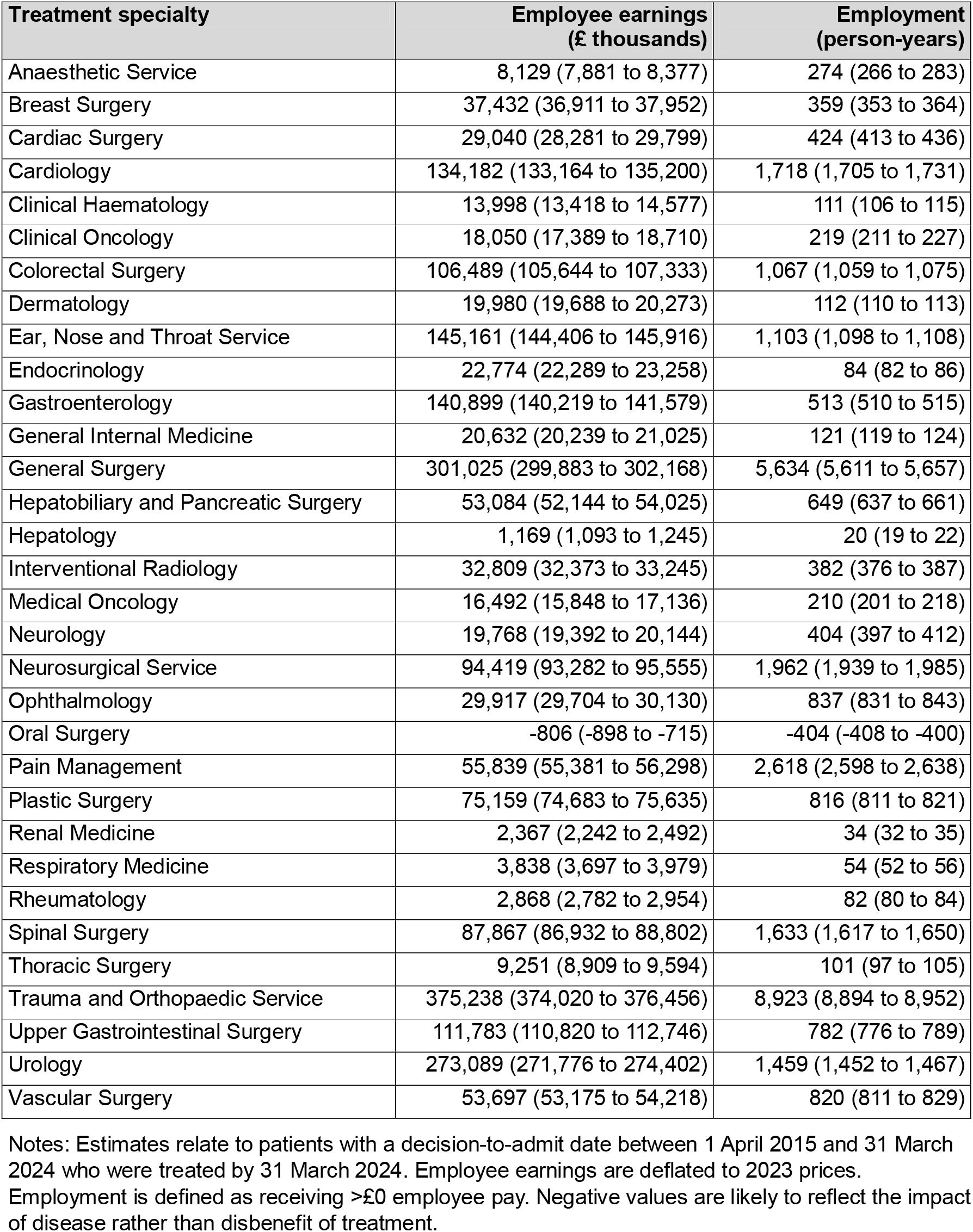
Estimated labour market impacts (point estimates and 95% confidence intervals) of above-target waiting times over the study period, accrued over five years post-admission, with target waiting times estimated from the gamma distribution.

| Treatment specialty | Employee earnings (£ thousands) | Employment (person-years) |
| --- | --- | --- |
| Anaesthetic Service | 8,129 (7,881 to 8,377) | 274 (266 to 283) |
| Breast Surgery | 37,432 (36,911 to 37,952) | 359 (353 to 364) |
| Cardiac Surgery | 29,040 (28,281 to 29,799) | 424 (413 to 436) |
| Cardiology | 134,182 (133,164 to 135,200) | 1,718 (1,705 to 1,731) |
| Clinical Haematology | 13,998 (13,418 to 14,577) | 111 (106 to 115) |
| Clinical Oncology | 18,050 (17,389 to 18,710) | 219 (211 to 227) |
| Colorectal Surgery | 106,489 (105,644 to 107,333) | 1,067 (1,059 to 1,075) |
| Dermatology | 19,980 (19,688 to 20,273) | 112 (110 to 113) |
| Ear, Nose and Throat Service | 145,161 (144,406 to 145,916) | 1,103 (1,098 to 1,108) |
| Endocrinology | 22,774 (22,289 to 23,258) | 84 (82 to 86) |
| Gastroenterology | 140,899 (140,219 to 141,579) | 513 (510 to 515) |
| General Internal Medicine | 20,632 (20,239 to 21,025) | 121 (119 to 124) |
| General Surgery | 301,025 (299,883 to 302,168) | 5,634 (5,611 to 5,657) |
| Hepatobiliary and Pancreatic Surgery | 53,084 (52,144 to 54,025) | 649 (637 to 661) |
| Hepatology | 1,169 (1,093 to 1,245) | 20 (19 to 22) |
| Interventional Radiology | 32,809 (32,373 to 33,245) | 382 (376 to 387) |
| Medical Oncology | 16,492 (15,848 to 17,136) | 210 (201 to 218) |
| Neurology | 19,768 (19,392 to 20,144) | 404 (397 to 412) |
| Neurosurgical Service | 94,419 (93,282 to 95,555) | 1,962 (1,939 to 1,985) |
| Ophthalmology | 29,917 (29,704 to 30,130) | 837 (831 to 843) |
| Oral Surgery | -806 (-898 to -715) | -404 (-408 to -400) |
| Pain Management | 55,839 (55,381 to 56,298) | 2,618 (2,598 to 2,638) |
| Plastic Surgery | 75,159 (74,683 to 75,635) | 816 (811 to 821) |
| Renal Medicine | 2,367 (2,242 to 2,492) | 34 (32 to 35) |
| Respiratory Medicine | 3,838 (3,697 to 3,979) | 54 (52 to 56) |
| Rheumatology | 2,868 (2,782 to 2,954) | 82 (80 to 84) |
| Spinal Surgery | 87,867 (86,932 to 88,802) | 1,633 (1,617 to 1,650) |
| Thoracic Surgery | 9,251 (8,909 to 9,594) | 101 (97 to 105) |
| Trauma and Orthopaedic Service | 375,238 (374,020 to 376,456) | 8,923 (8,894 to 8,952) |
| Upper Gastrointestinal Surgery | 111,783 (110,820 to 112,746) | 782 (776 to 789) |
| Urology | 273,089 (271,776 to 274,402) | 1,459 (1,452 to 1,467) |
| Vascular Surgery | 53,697 (53,175 to 54,218) | 820 (811 to 829) |

Notes: Estimates relate to patients with a decision-to-admit date between 1 April 2015 and 31 March 2024 who were treated by 31 March 2024. Employee earnings are deflated to 2023 prices. Employment is defined as receiving >£0 employee pay. Negative values are likely to reflect the impact of disease rather than disbenefit of treatment.

**Supplementary Figure 1.**
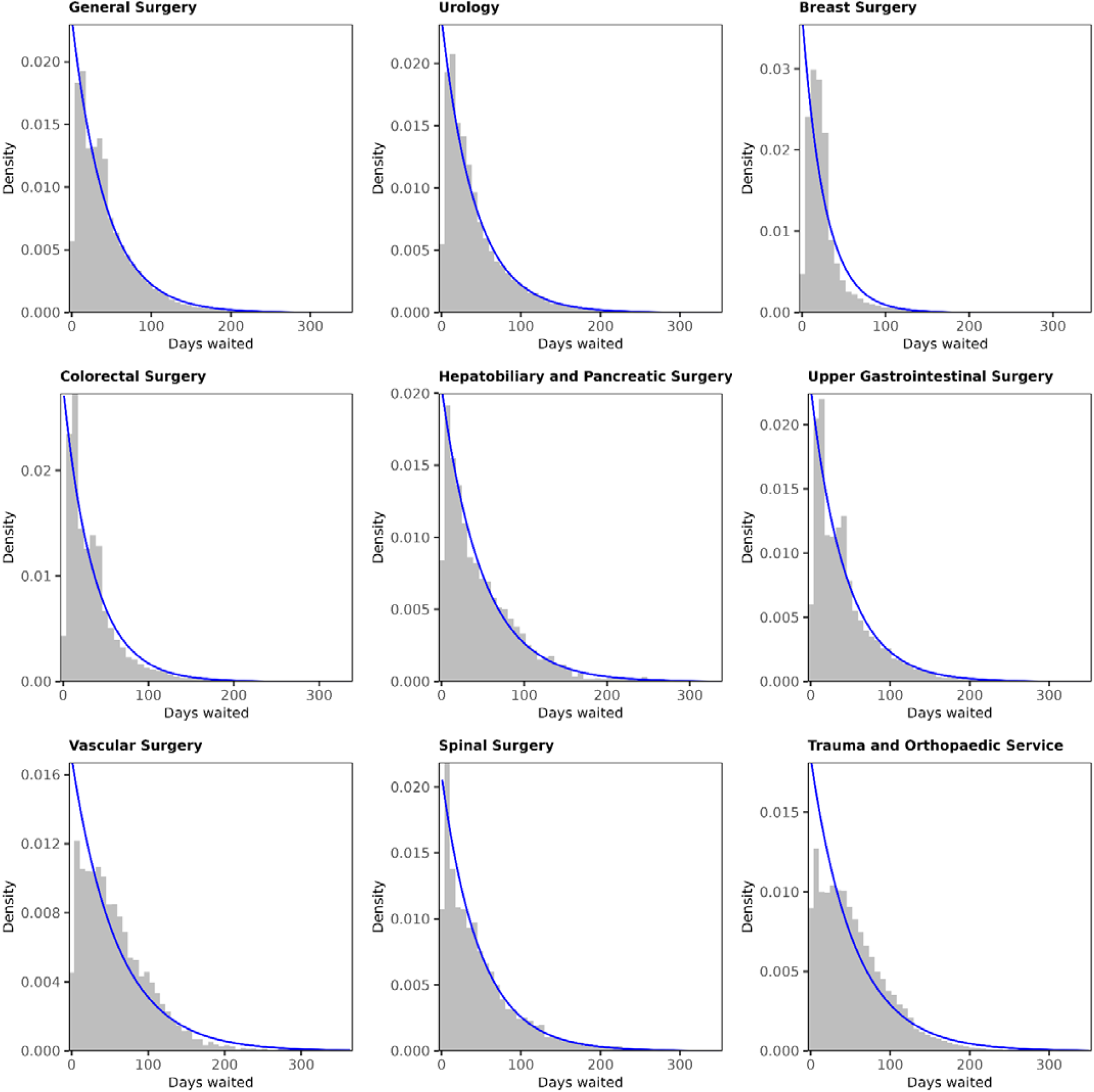
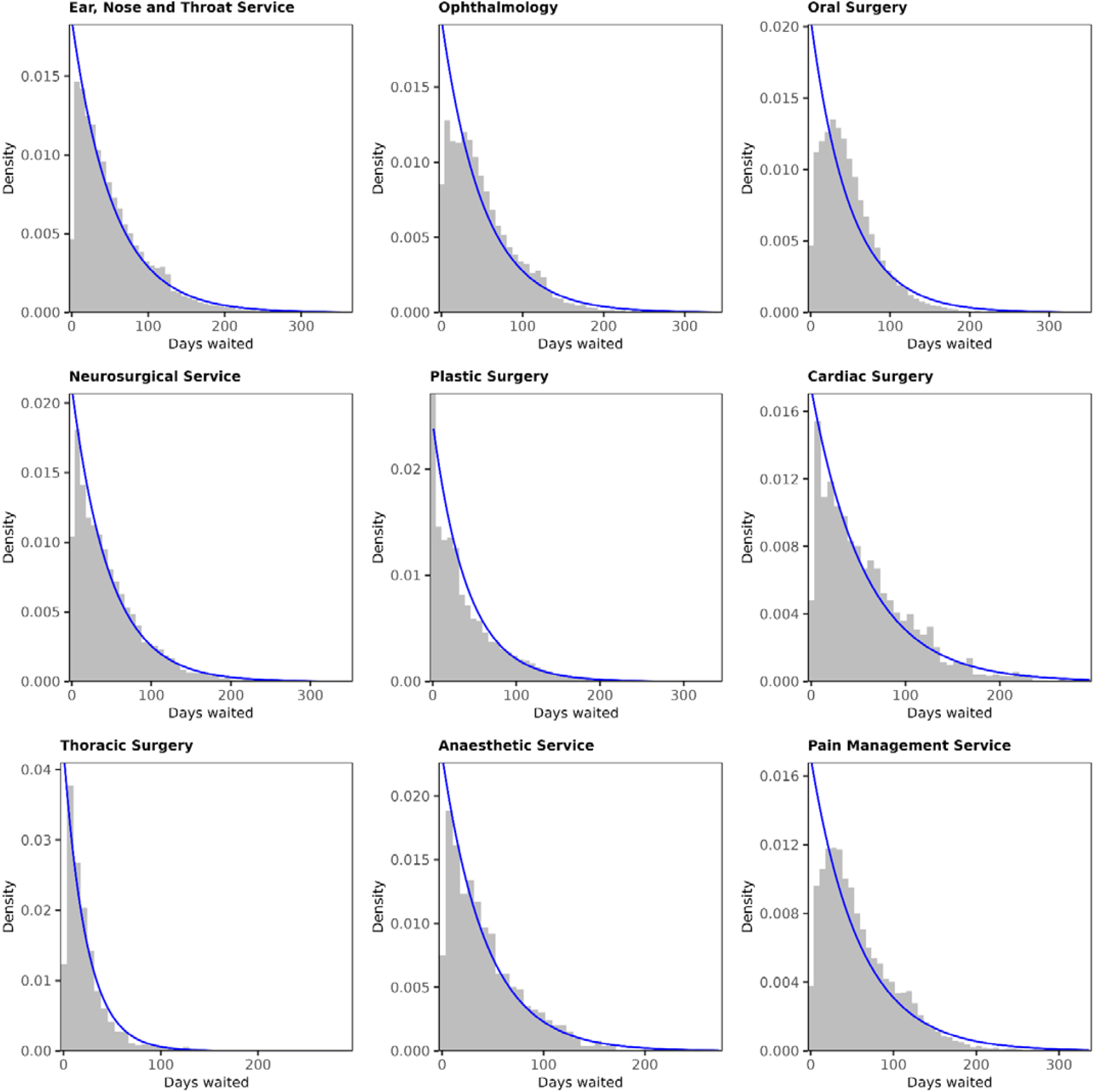
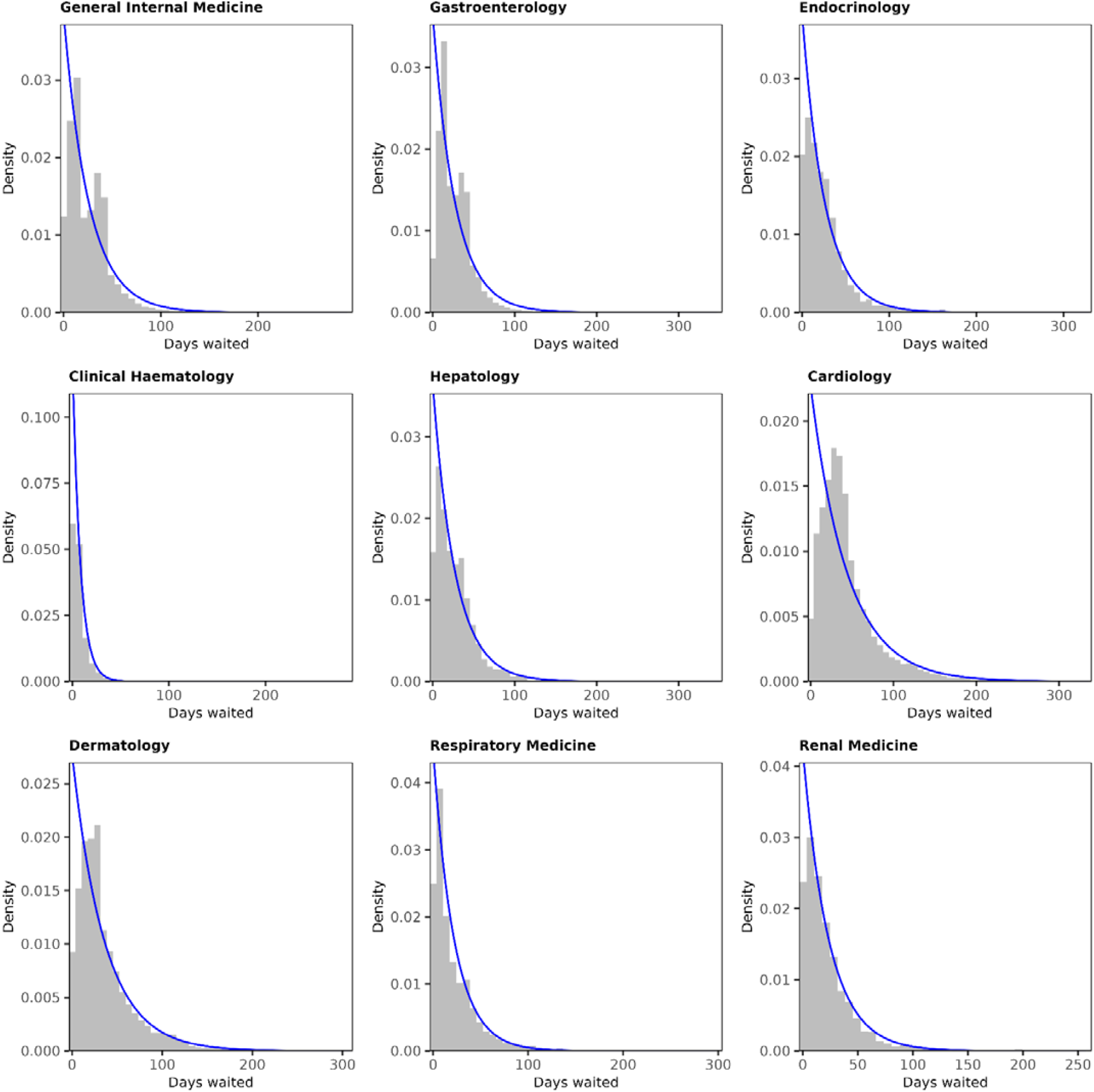
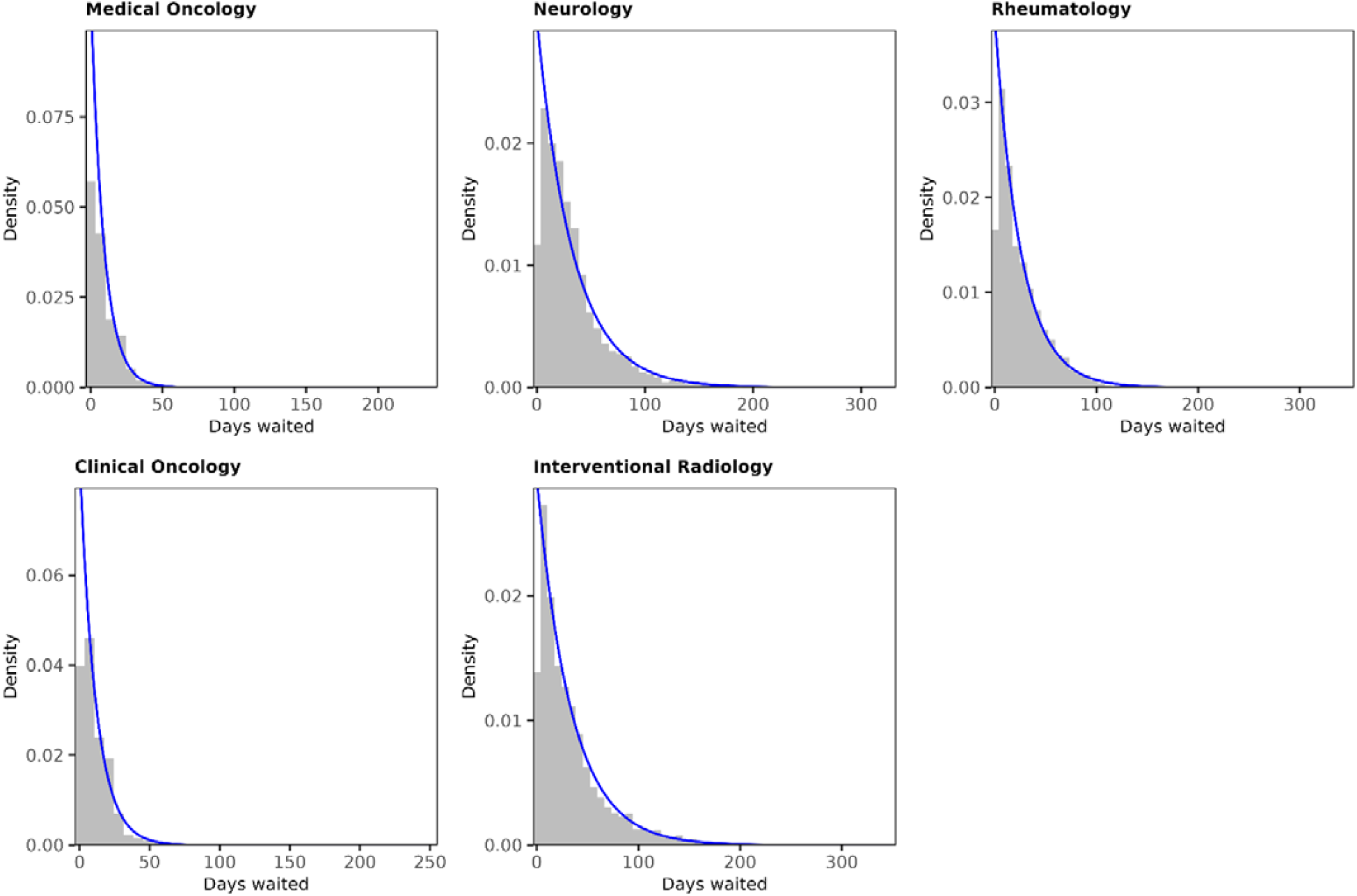
Histograms of waiting times for admissions in 2015/16 with the fitted exponential probability density function overlaid

**Supplementary Figure 2.**
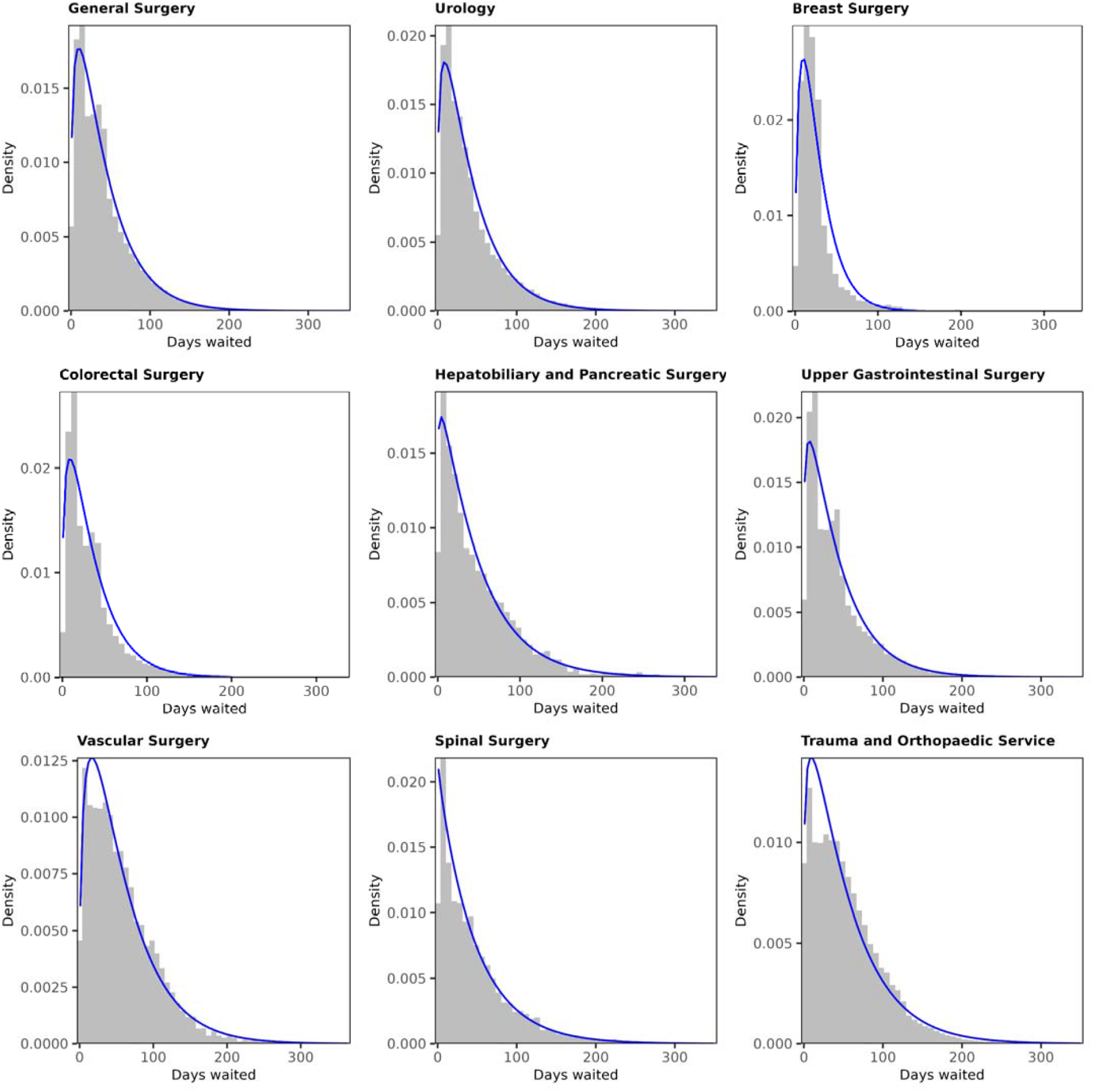
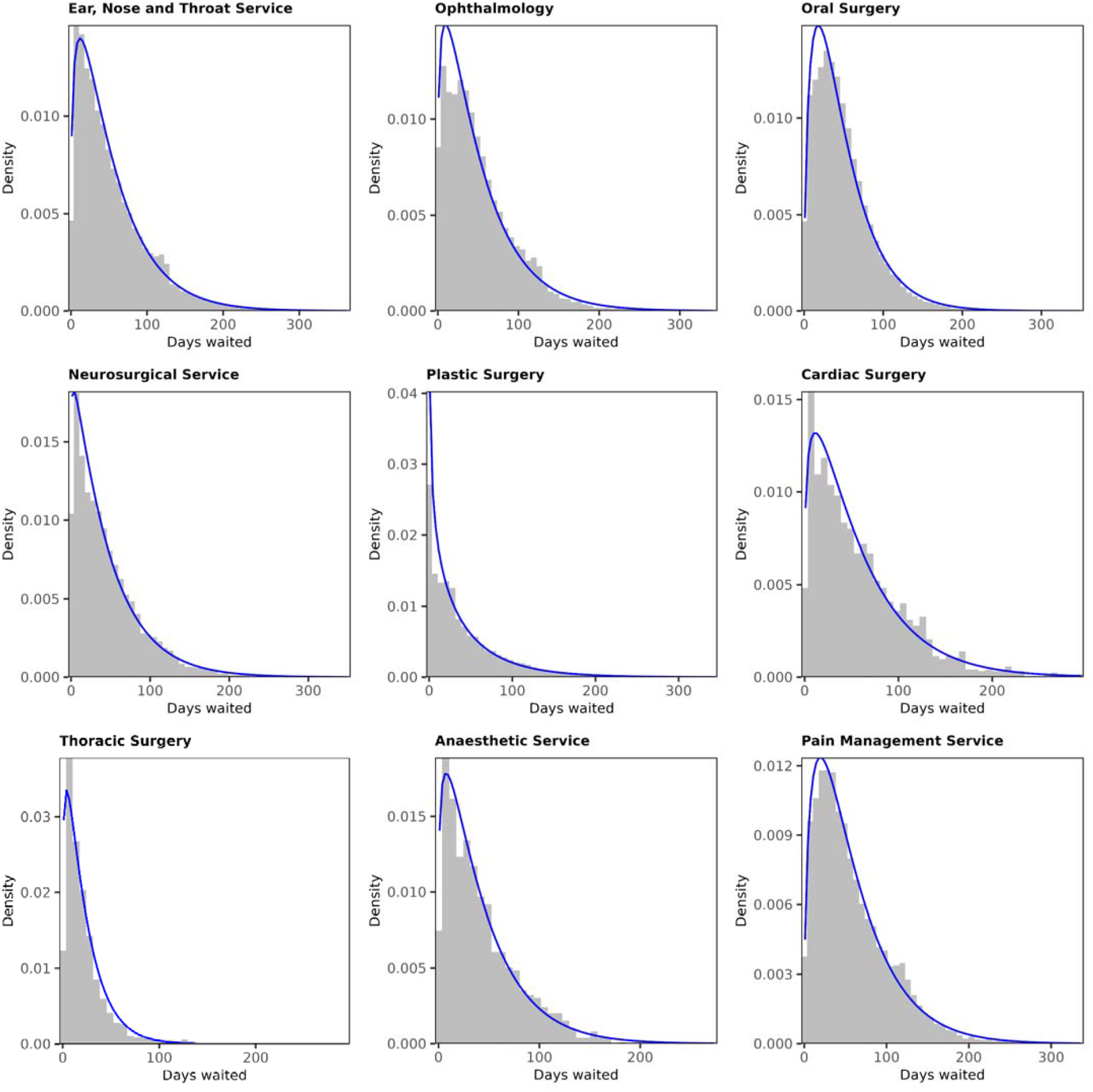
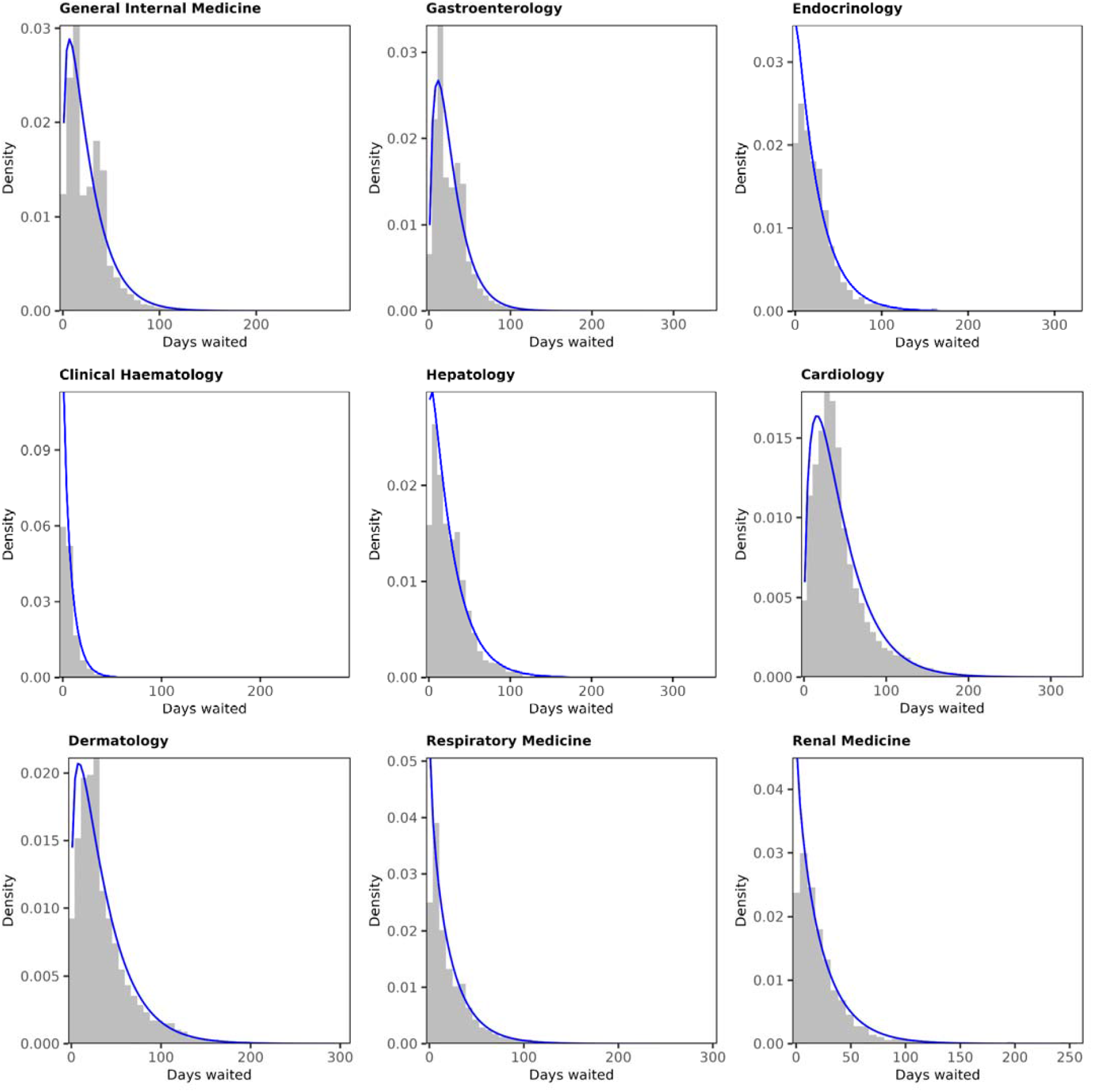
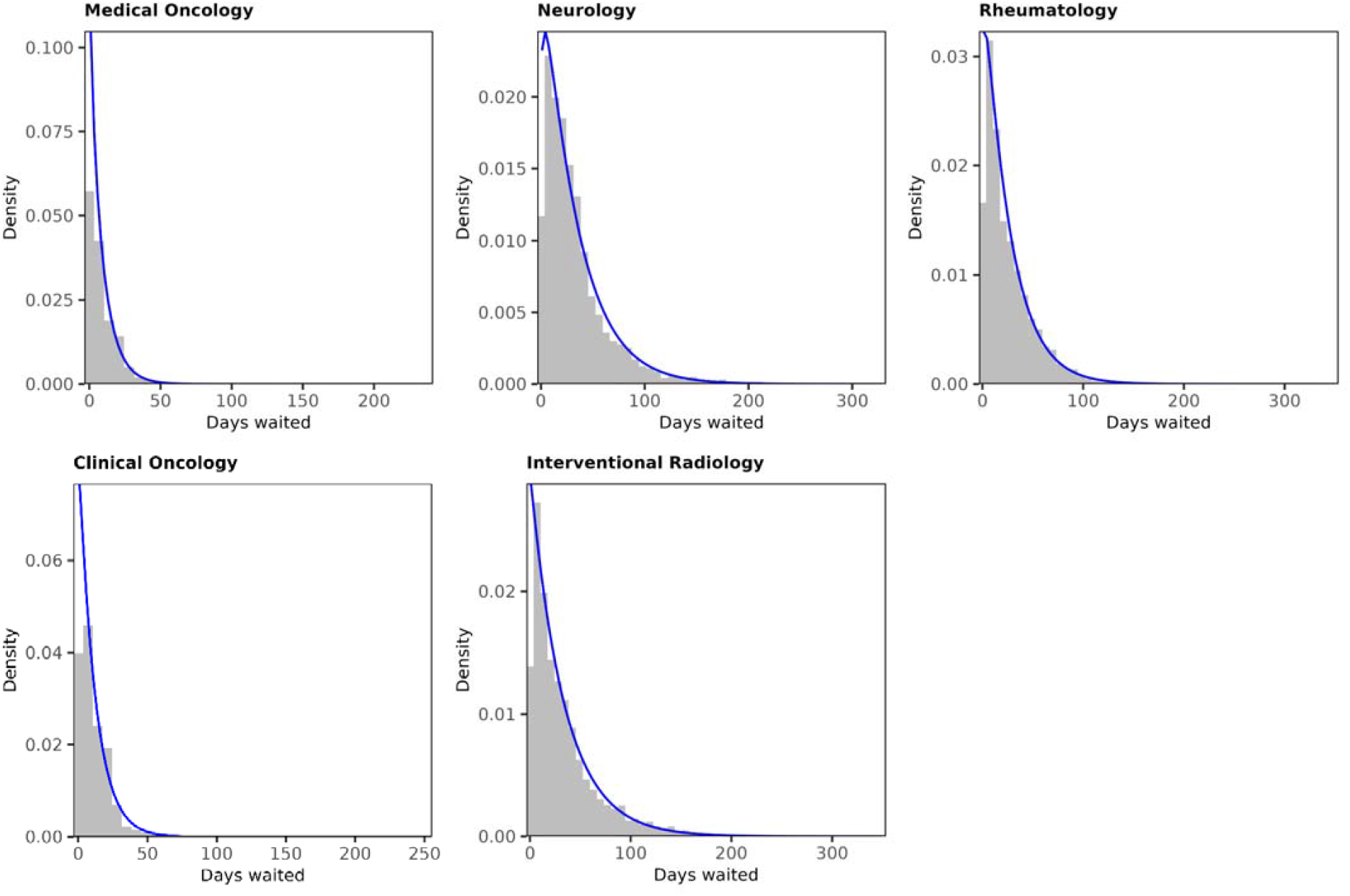
Histograms of waiting times for admissions in 2015/16 with the fitted gamma probability density function overlaid

## Supplementary Appendix A: Detailed statistical methodology

### Calculating target waiting times

For each treatment specialty (*j*) in each financial year of the study period (*r*), we calculated each patient’s percentile (*p*_*i*_) of an exponential distribution with the rate parameter (*λ*) estimated as the inverse of the mean waiting time 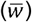:

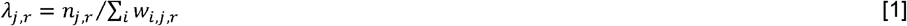

where *n*_*j,r*_ is the number of patients treated in specialty *j* in year *r*. For each treatment specialty, the target waiting time distribution was an exponential distribution with a rate parameter estimated as the inverse of the mean waiting time in 2015/16, calculated from the observed HES data:

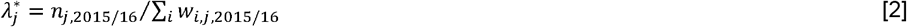

Each patient’s target waiting time 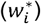 was then taken from the target distribution (defined by rate parameter *λ*^*^), had they remained at the same percentile of the observed waiting time distribution (*p*_*i*_ ).

Given that average waiting times were lower in 2015/16 than in other years of the study period, the entire distribution was shifted to the left when moving from observed to target waiting times, with those in the upper tail of the distribution experiencing the biggest changes. By way of example, this is illustrated in **Figure A1** for elective Trauma and Orthopaedic Service inpatients, where the median waiting time fell from 57 days (observed for 2015/16 to 2023/24 admissions) to 40 days (target based on 2015/16 admissions).

**Figure A1.**
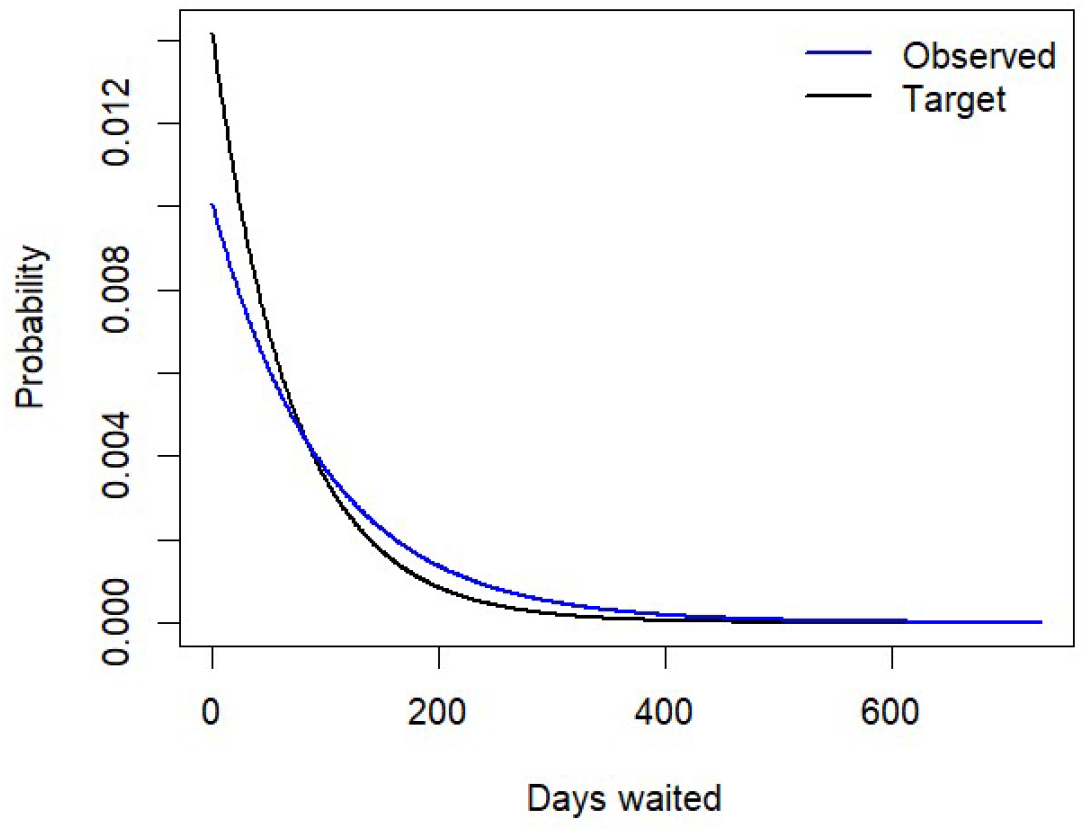
Exponential probability density functions for observed (2015/16 to 2023/24) and target (2015/16) waiting times, elective Trauma and Orthopaedic Service inpatients

To align with the exposure and treatment effects described below – which are expressed in terms of month before and after treatment, respectively – the observed and target waiting times were rounded from days to calendar months when performing all subsequent calculations.

### Quantifying the labour market cost of reduced waiting times

#### Cost of extended waiting times, component 1: less treatment effect accrued

We used published estimates of the labour market treatment effects (in terms of employee earnings and person-years of employment) associated with elective inpatient procedures^1^, the methodology for which has been detailed elsewhere^2^. In summary, the Hospital Episode Statistics (HES) Admitted Patient Care (APC) dataset was linked to Pay-As-You-Earn (PAYE) tax records from His Majesty’s Revenue and Customs (HMRC) to estimate monthly average employee pay (deflated to 2023 prices) and the monthly probability of paid employment, for months (*t*) before and after first elective inpatient admission within each treatment specialty. The monthly values (*y*_*j,t*_) were age-standardised 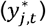 to the half-year age distribution in the month of treatment.

A segmented linear regression model was fitted to 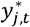 over the 12 months prior to treatment to infer onset of illness, identified by a breakpoint in the pay/employment trajectory:

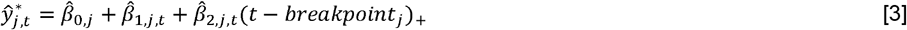

where( *t* − *breakpoint*_*j*_)_+_ = (*t* − *breakpoint*_*j*_) if *t* > *breakpoint*_*j*_ and 0 otherwise; 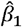 represents the slope of the pre-breakpoint trajectory; and 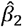 represents the change in slope after the breakpoint.

The model was used to extrapolate the trajectory from the post-onset, pre-treatment epoch over the post-treatment period. The treatment effect was then estimated as the difference between the observed and model-projected values of age-standardised pay/employment in each month up to five years following treatment:

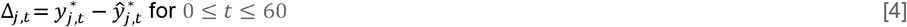

The projected values represent a counterfactual outcome; that is, the expected trajectory of age-standardised pay/employment had individuals not received treatment and instead remained in their ‘pre-treatment’ state. This interrupted time series (ITS) design is illustrated in **Figure A2**.

**Figure A2.**
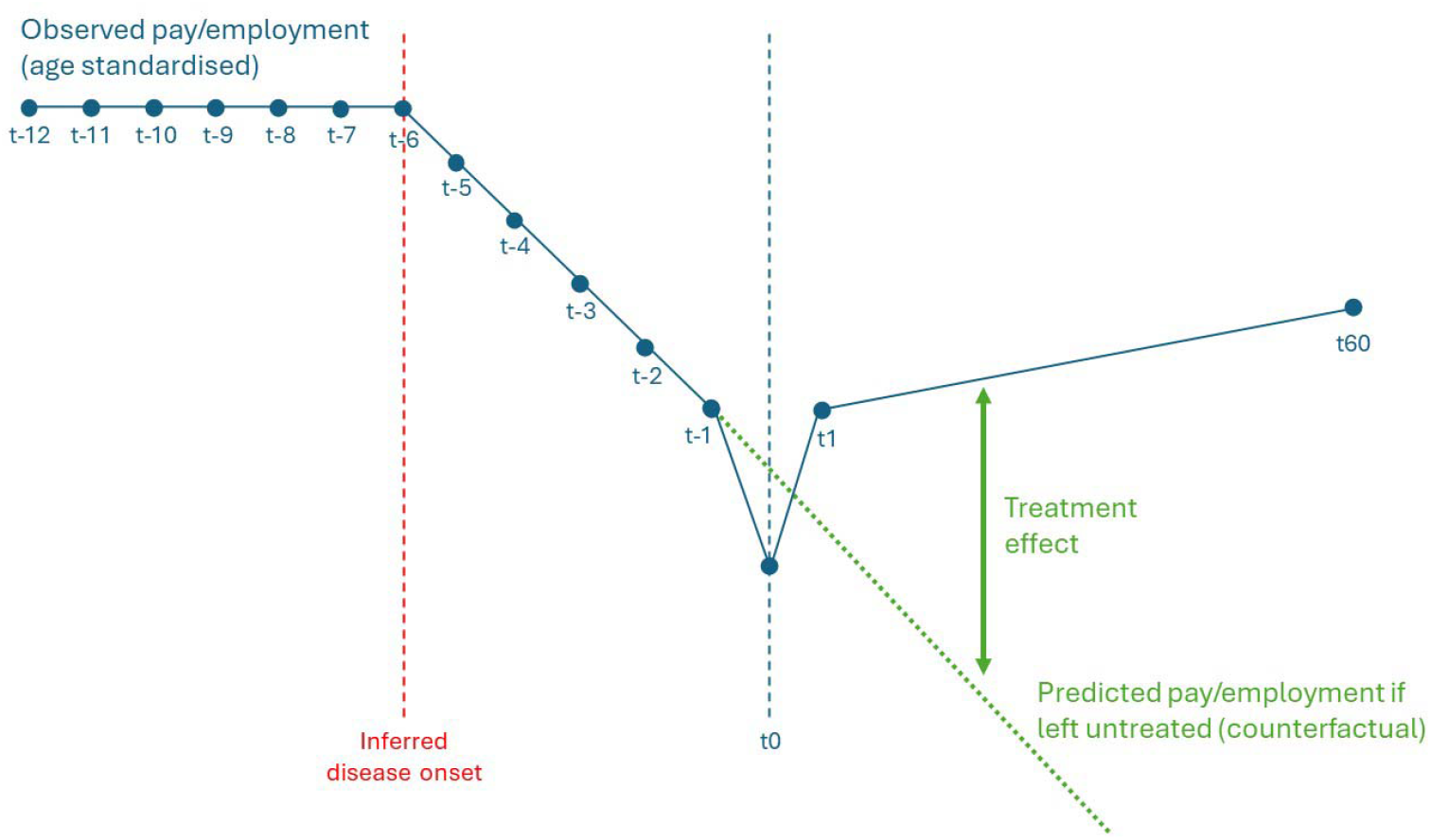
Stylised illustration of how treatment effects are estimated

We aimed to estimate how much treatment effect (Δ_*j,t*_) would be accrued within an evaluation period of five years from decision-to-admit under target waiting times, compared with what was actually accrued given the observed waiting times over the study period. We considered two different measures of treatment effect (each relative to the counterfactual of no treatment):

- Additional earnings accrued over the five-year evaluation period
- Additional person-months of employment accrued over the five-year evaluation period

For example, if a patient waited three months for treatment, they accrued treatment effects from Month 0 to Month 57. However, had their waiting time been reduced to one month, they would have accrued treatment effects from Month 0 to Month 59. In general, each patient accrued 60 − *w*_*i*_ months of treatment effect given observed waiting times, and 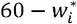 months of treatment effect under target waiting times (where 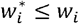). This notion of accruing additional treatment effect over the five-year period from decision-to-admit had waiting times been at their target levels is illustrated in **Figure A3**.

**Figure A3.**
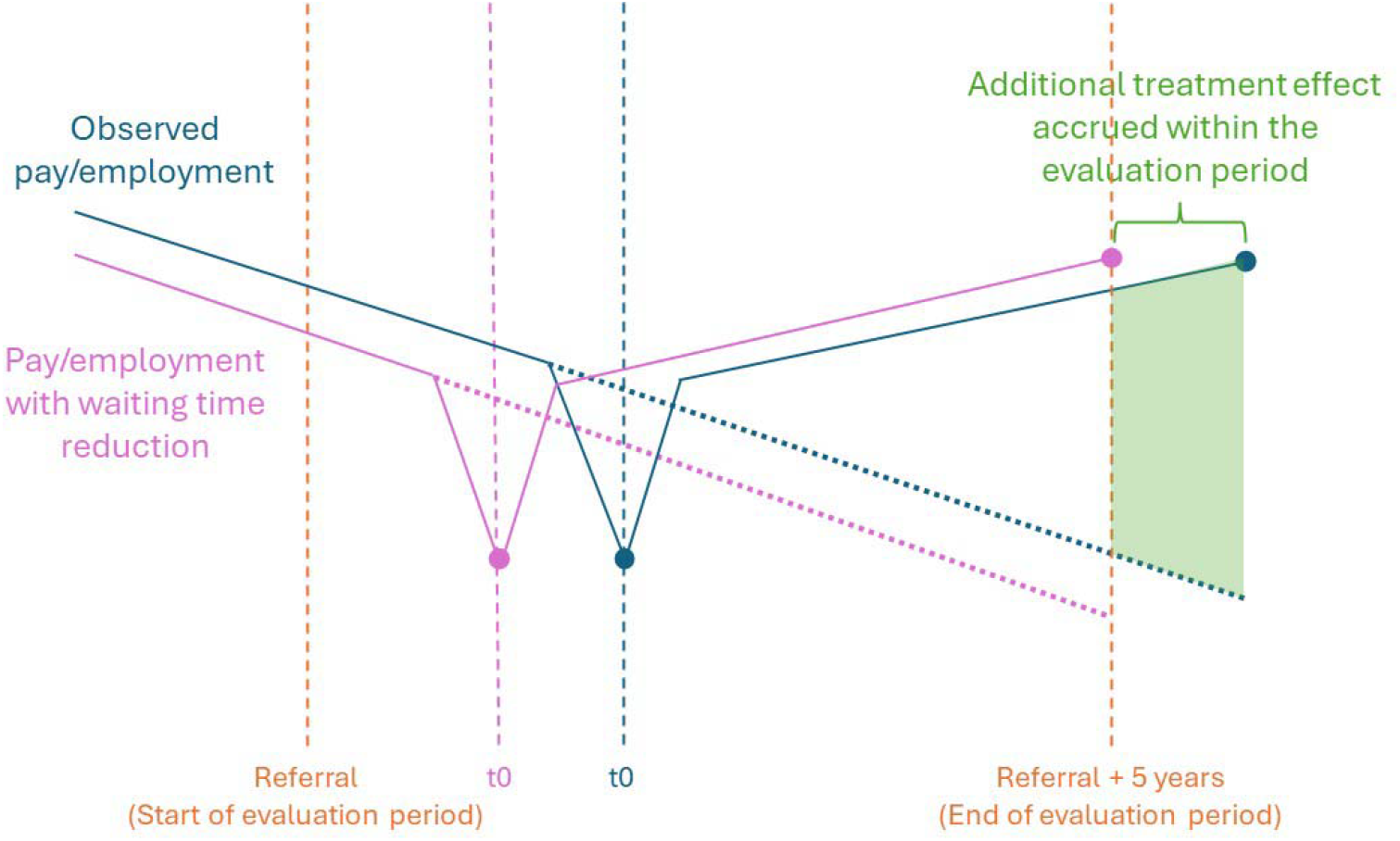
Stylised illustration of accruing additional treatment effect over the five-year period from decision-to-admit had waiting times been at their target levels

#### Cost of extended waiting times, component 2: more disbenefit of disease accrued

We fitted the same segmented linear regression model as outlined in equation [3] to the published age-standardised pay/employment trajectories^1^ to estimate the effect of exposure to disease prior to treatment. The pay/employment trajectory from the ‘pre-disease’ epoch (from Month -12 to the breakpoint) was projected over the ‘pre-treatment’ epoch (from the breakpoint to Month -1) to provide a counterfactual outcome: an estimate of individuals’ pay/employment had they remained in the ‘pre-disease’ state, as illustrated in **Figure A4**.

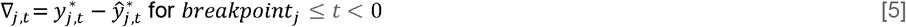

**Figure A4.**
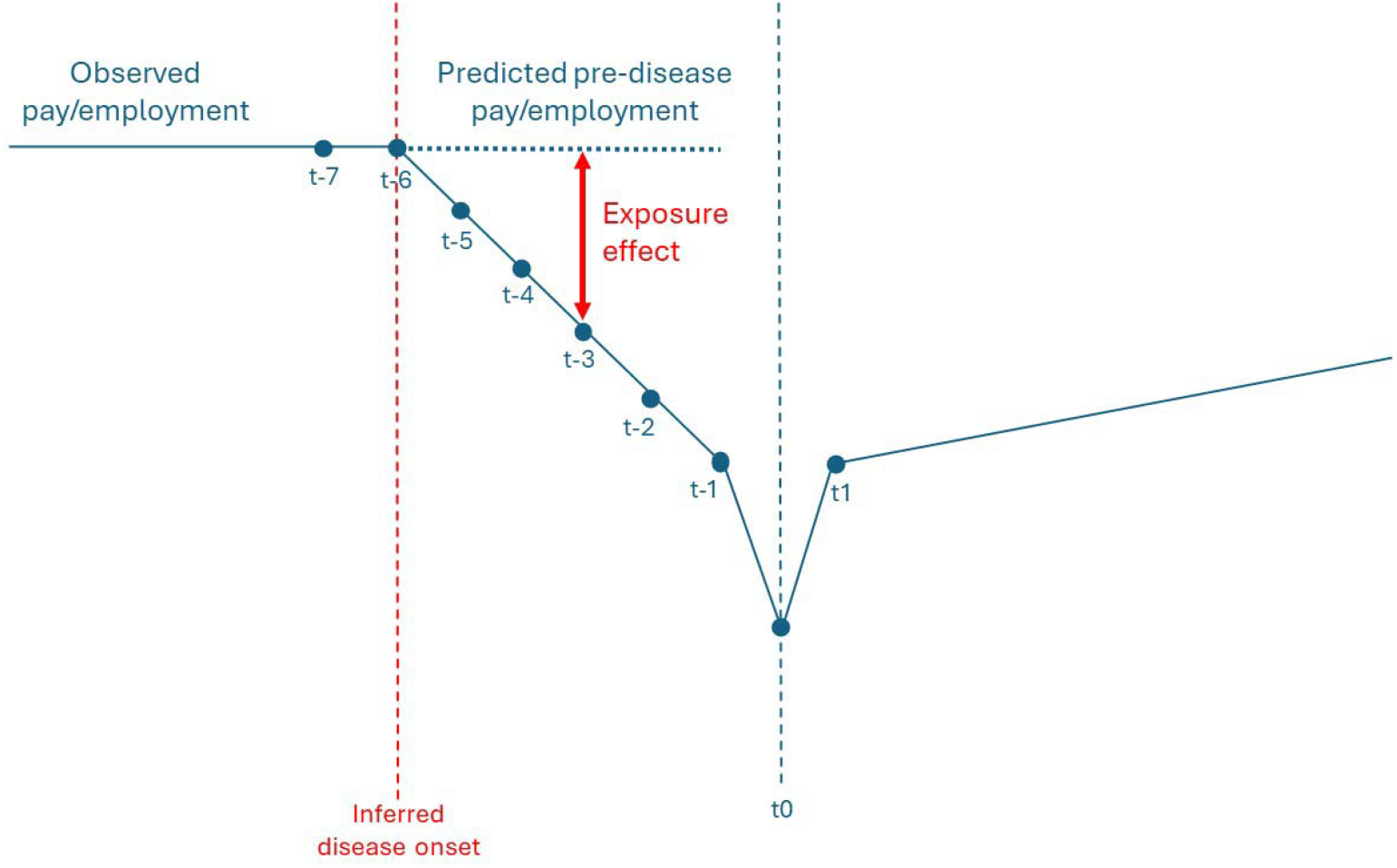
Stylised illustration of how exposure effects are estimated

When waiting times are above their target levels, patients spend more time in the ‘pre-treatment’ epoch, thus they accrue more disbenefit of illness in terms of their pay/employment (∇_*j,t*_).

For example, if a patient spent five months in the ‘pre-treatment’ epoch, they accrued five months of disbenefit due to illness (from Month -5 to Month -1). However, had their waiting time been one month shorter, they would have accrued just four months of disbenefit (from Month -5 to Month -2). In general, if patients’ observed waiting times exceed their target waiting times by *x*_*i*_ months (i.e. 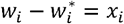), they would accrue additional disbenefit of illness from Month -1 to Month -*x*_*i*_, where Month -*x*_*i*_ was capped at the breakpoint. This is illustrated in **Figure A5**.

**Figure A5.**
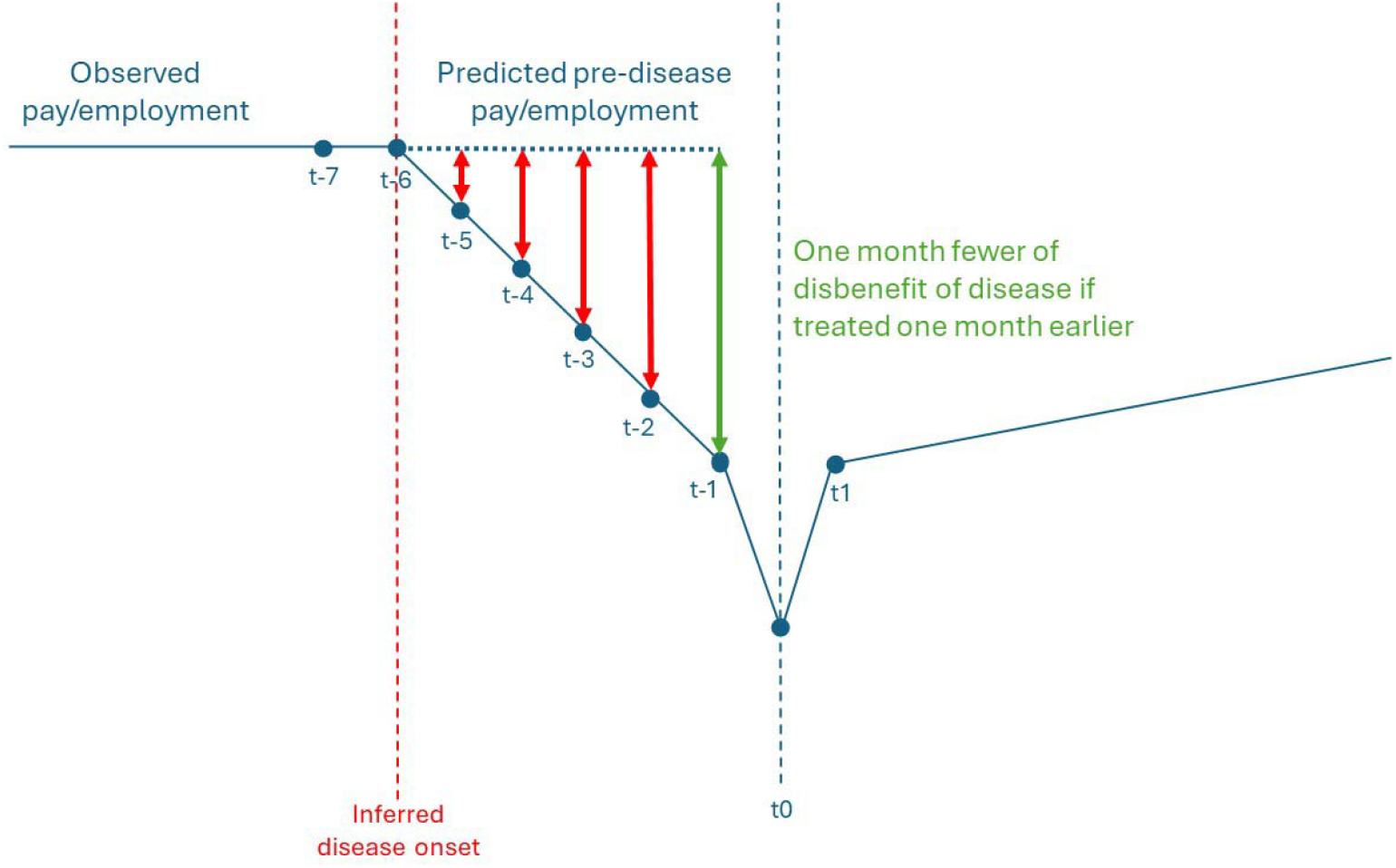
Stylised illustration of accruing less disbenefit of illness in the ‘pre-treatment’ epoch had waiting times been at their target levels

#### Cost of extended waiting times, component 3: more pre-treatment health deterioration so pay/employment progresses to a lower level following treatment

When waiting times are above target, patients progress further along the pay/employment trajectory in the ‘pre-treatment’ epoch than they otherwise would have done. Therefore, the post-treatment turning point in the trajectory occurs at a lower level of pay/employment (assuming a downward pre-treatment trajectory), and this difference in level was assumed to be constant and to persist throughout the post-treatment period. As the counterfactual pay/employment trajectory was assumed to be unaffected, the constant that was added to the observed pay/employment trajectory in each month after treatment translated to the same constant added to the treatment effect in each month.

For example, if a patient was treated one month earlier than observed, the change in the pay/employment trajectory that occurred between Month -1 and Month 0 would actually have begun in Month -2. Thus, the difference in level between Month -1 and Month -2 was added as a constant to the observed trajectory (and the treatment effect) from Month 0 to Month 60. In general, if patients’ observed waiting times exceeded their target times by *x* _*i*_ months, the constant added to pay/employment in each month post-treatment was equal to the difference in pay/employment between Month -1 and Month [-1-*x*], where Month [-1-*x*] was capped at the breakpoint. This is illustrated in **Figure A6**.

**Figure A6.**
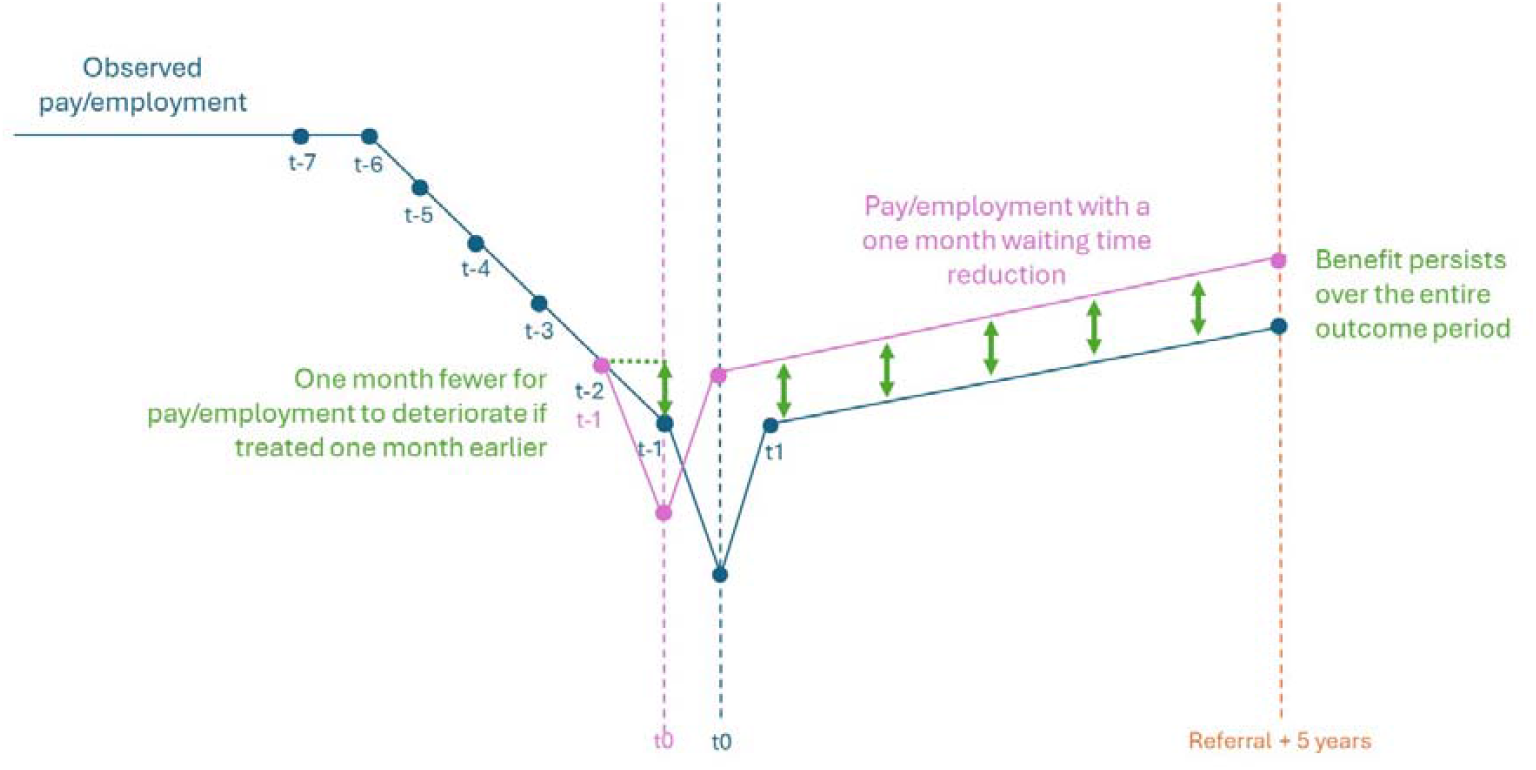
Stylised illustration of the effect of less pre-treatment health deterioration on post-treatment outcomes had waiting times been at their target levels

### Total labour market losses of extended waiting times

For each treatment specialty, the change in the cumulative treatment effect over the evaluation period following decision-to-admit, plus the disbenefit of more exposure to disease, due to waiting times being above their target levels was calculated as follows:

[Cumulative treatment effect given target waiting times

−

Cumulative treatment effect given observed waiting times]

+

Effect of accruing more disbenefit of disease

+

Effect of more pre-treatment health deterioration on post-treatment levels

As the 32 study cohorts are not mutually exclusive, some of the estimated loss in pay/employment due to above-target waiting times 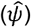 for a given treatment specialty may be attributable to patients being treated in other specialities. Thus, there will be some degree of double-counting if estimates of the labour market costs of extended waiting times are simply summed across treatment specialties, giving an upper bound on the estimated total loss due to above-target waits:

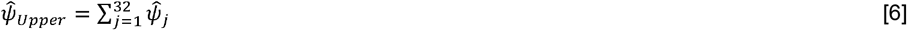

To provide a more conservative estimate, cohort sizes (*n*_*j*_) were re-derived 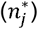 under the assumption that patients belonging to multiple study cohorts experienced losses in pay/employment due to above-target waiting times for just one specialty in which they were treated: the one with the smallest per-person effect size. The re-calculated cohort sizes were then multiplied by the per-person effect sizes and summed across the 32 treatment specialties, giving a lower bound on the estimated total loss due to above-target waits:

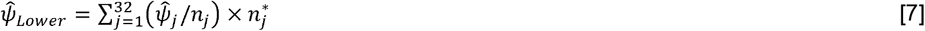

For each treatment specialty, 95% confidence intervals were constructed around the point estimate of the total labour market loss of extended waiting times by assuming that this quantity follows a normal distribution:

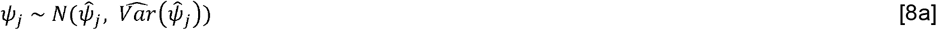

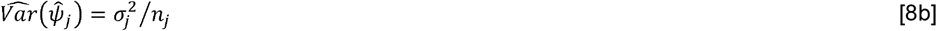

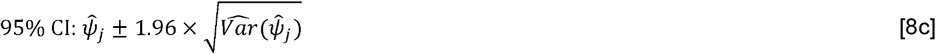

where *σ*^2^ is the sample variance of *ψ* among all patients treated within the treatment specialty. The data were aggregated from person-months to persons when calculating the confidence intervals. Confidence intervals around 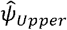 and 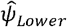 were derived by using the lower and upper limits of the confidence intervals around 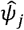 in place of 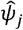 in equations [6] and [7].

